# Pain and Pain Sensitivity Assessments in the Acute to Chronic Pain Signatures (A2CPS) Program

**DOI:** 10.64898/2026.08.14.26360457

**Authors:** Laura A. Frey-Law, Giovanni Berardi, Briha Ansari, Yanxi Liu, Bella Satpathy-Horton, Kathleen A. Sluka, Carol GT Vance, Dana L Dailey, Robert J. McCarthy, Tor D. Wager, Martin A. Lindquist, Steven E. Harte, the A2CPS Consortium

## Abstract

The Acute to Chronic Pain Signatures (A2CPS) project is a large, multisite, longitudinal observational study designed to identify biomarkers that predict the transition from acute to chronic pain following surgery in more than 2200 patients. Two participant cohorts were recruited before undergoing either knee arthroplasty or thoracic surgery. A unique feature of this study is its comprehensive evaluation of pain, including evoked and recall pain measures collected at baseline, 6-weeks, and 3-months following surgery, in addition to the primary pain outcome assessed remotely at 6 months. This paper describes the acquisition, quality control procedures, and available pain and pain sensitivity variables included in the A2CPS study. Self-report pain assessments include surgical site (i.e., index) pain intensity, pain interference and quality, spatial distribution of pain using body maps, and pain-related dysfunction specific to each cohort. Quantitative sensory testing yielded evoked pain sensitivity data including pressure pain thresholds, temporal summation of pain, dynamic mechanical allodynia, and conditioned pain modulation at both index and common sites across cohorts. Movement-evoked pain was assessed for each cohort using relevant functional tasks (knee: 10m walk and five-time-sit-to-stand tests, thoracic: deep breathing and coughing). Using baseline data from release v2.1.0, comprising approximately 1,400 participants, we evaluated interrelationships among pain variables. Overall, the A2CPS pain and pain sensitivity data provide a robust, comprehensive set of variables that supports the study’s goal of uncovering predictive biomarkers of post-operative chronic pain and enables broader exploration relative to other study outcomes, including imaging, psychosocial, and “omics” data.

## Introduction

Pain, particularly persistent and chronic pain, affects roughly one-quarter of U.S. adults, with nearly 10% living with high-impact chronic pain^1–3^. Unlike acute pain, which often serves a protective role, chronic pain persists beyond normal healing time and often lacks a clear underlying mechanism, making targeted treatment challenging. Chronic pain is associated with reduced quality of life, increased risk of anxiety and depression, and substantial economic costs – exceeding $600 billion annually in healthcare costs and lost productivity^4,5^ – underscoring its status as a major public health concern.

The Acute to Chronic Pain Signatures (A2CPS) program, funded by the NIH Common Fund, is a large, multisite, longitudinal observational study designed to identify predictive biomarkers of chronic pain, 6-months following surgery. A2CPS incorporated a comprehensive, multidimensional battery of pain and pain sensitivity measures, often lacking in longitudinal studies. The study recruited over 2200 patients undergoing either knee arthroplasty or thoracic surgery. Self-report and evoked-pain assessments were collected across baseline (pre-operatively) and follow-up visits at 6 weeks, 3 months, and 6 months post-operatively, although the specific assessments administered varied by timepoint. All testing was conducted at six clinical sites despite broader recruitment across participating hospitals. This rich pain dataset supports the study’s primary goal of uncovering predictive biomarkers of chronic postsurgical pain and enables more nuanced exploration of pain responses relative to other study domains, including imaging, psychosocial, and omics data.

Pain is a complex construct that requires a multidimensional framework to capture its sensory, affective, cognitive, behavioral, and sociocultural dimensions^6–10^. Although numerous biopsychosocial factors have been implicated as potential biomarkers, no single marker fully explains the pain experience or predicts chronic pain development. The A2CPS protocol was designed to incorporate multiple pain domains, such as pain at rest and during movement, pain interference, widespread pain, and pain sensitivity using quantitative sensory testing (QST), both as pre-operative predictors and as indicators of post-operative response at several time points. In addition to assessments at 6 weeks and 3 months, daily pain ratings collected from 3 to 28 days after surgery provide a detailed characterization of the early post-operative pain trajectory. The final pain outcomes rely on self-report assessments at 6-months, with repeated assessments over 7 days allowing for exploration of their temporal stability. Finally, there is a voluntary pain assessment at 12 months.

This manuscript details the acquisition, quality control, and availability of pain and pain sensitivity variables in the A2CPS dataset. We also characterize inter-relationships among baseline pain variables using correlations, factor analysis, and dynamic linkage models to illustrate the multidimensional structure of these pain constructs. By presenting these baseline data and their empirical structure within this series of A2CPS consortium papers, we aim to facilitate informed planning and interpretation of future secondary analyses of the dataset by the broader research community.

## Methods

This manuscript provides a summary of baseline (pre-operative) pain and pain sensitivity data from the A2CPS Consortium (Data Release 2.1.0). All participants provided electronic informed consent as approved by the central Institutional Review Board housed at the University of Iowa.

### Self-Report Pain Assessments

Multiple validated self-report assessments were completed at baseline, daily during the acute post-operative period (days 3-28), and at 6-weeks, 3-months, and 6-months following surgery, with an optional 12-month assessment also available^11,12^. See **Table 1** for a list of self-report pain metrics and their corresponding abbreviations, brief summaries and dataset variables available to represent or generate each assessment.

**Table 1.**
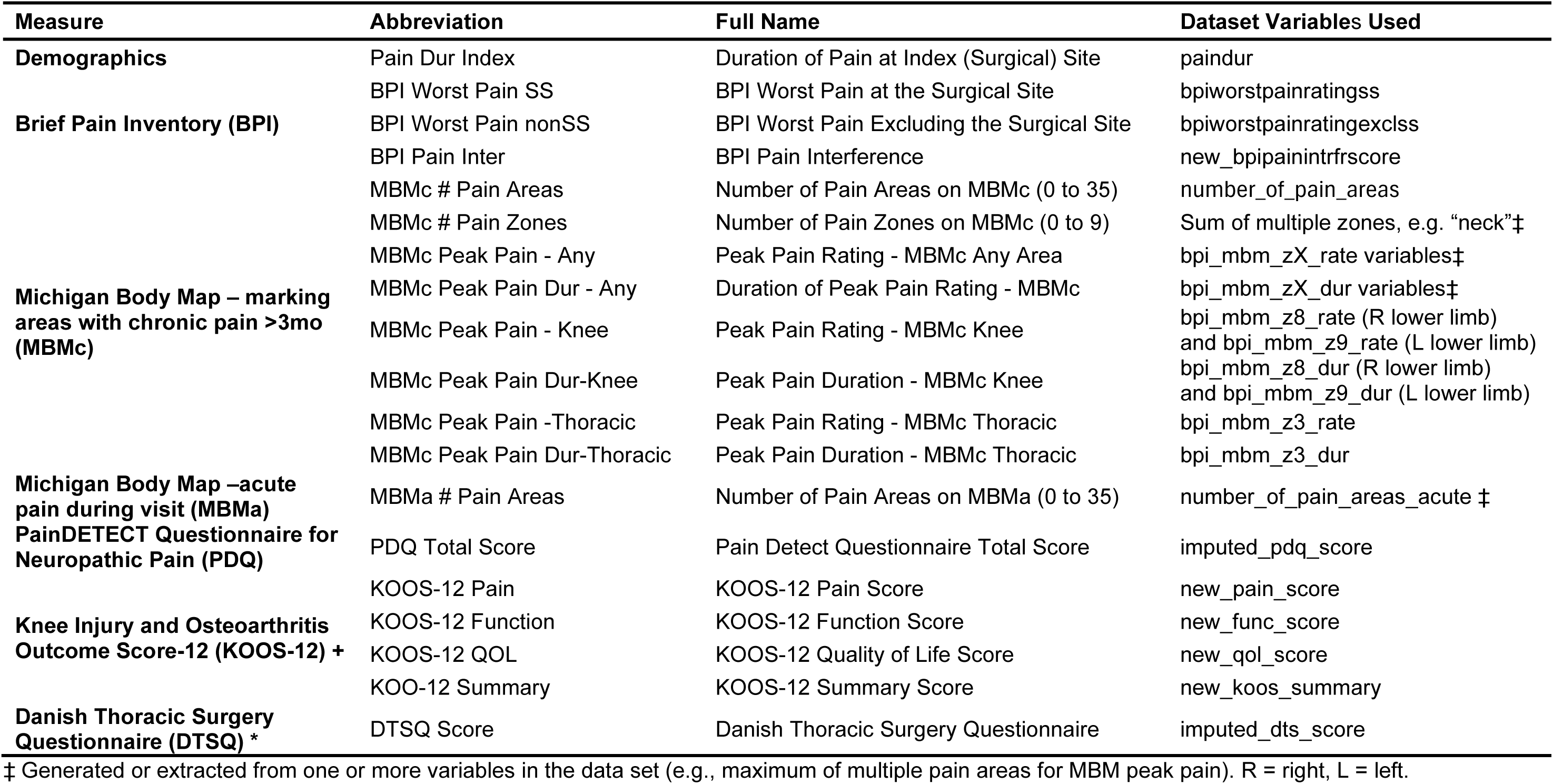
Self-report Pain Variables - List of Abbreviations.

#### Brief Pain Inventory - Worst Pain

The Brief Pain Inventory (BPI) is a commonly used tool to assess the nature of pain and its impact. The first part of the BPI, the pain severity scale, typically measures pain severity including worst, least, average, and current pain over the past 24 hours using an 11-point numeric rating scale (NRS), anchored by 0 representing "no pain" and 10 representing "pain as bad as you can imagine". To reduce subject burden and increase specificity, we modified these pain intensity items to assess only worst pain specifically at the surgical site (“knee” or “chest”) and separately for worst pain excluding the surgical region. The second part of the BPI, the pain interference scale (BPI PI), was used to assess how pain interferes with general activity, mood, walking, normal work, relationships with others, sleep, and enjoyment of life. Each item was ranked on an 11-point NRS, where 0 represents "does not interfere" and 10 denotes "completely interferes". The pain interference score was calculated as the sum of the pain interference items^13^.

#### Michigan Body Map (acute and chronic)

The Michigan Body Map (MBM) was used to assess the spatial extent of chronic pain (i.e., instructions indicated pain lasting 3 months or more)^14,15^. The MBM has 35 distinct body sites/areas participants may endorse (i.e., total area score ranges from 0 – 35). These 35 areas were grouped into 9 zones. When pain was endorsed for a zone, follow-up items included average pain intensity over the past 24 hours (0-10 NRS), and pain duration (< 1 month; 1 month or more, but < 6 months; 6 months or more, but < 2 years; or 2+ years). Recent evidence suggests that widespread body pain limits quality of life, impairs function, and is linked to altered pain processing within the central nervous system as well as genetic and immune system pathobiology^4–21^. Additionally, individuals with widespread pain often report co-occurring symptoms of fatigue, sleep dysfunction, and cognitive difficulty, which all contribute to limited treatment responsiveness^22–24^. In addition to the standard chronic pain version of the MBM, an acute pain version of the MBM was used to assess the distribution of current pain at the baseline and 3-month visits, without follow-up items.

#### Pain Trajectory Survey

To capture the time-varying aspects of pain and related symptoms, daily trajectory assessments were obtained only post-operatively days 3 to 28 and for 7 days at the 6 month follow-up, corresponding to the primary study outcome. Trajectory assessment is supported by prior research suggesting increased reliability and sensitivity of pain intensity measurement when using multiple assessment timepoints^25–27^. Among the items assessed, worst surgical site pain, average surgical site pain, and pain interference were measured over the past 24 hours using a 0-10 NRS. In addition, over-the-counter and prescription pain medication use for surgical site pain was also captured with dichotomous (yes/no) responses. These daily assessments enabled characterization of the pain trajectories acutely and at the 6 month endpoint.

#### PainDETECT

The painDETECT questionnaire (PDQ) screens for neuropathic components of pain^28,29^ and has been linked to worse pain and functional outcomes^30,31^. The first seven of nine questions assess the strength of several neuropathic pain symptoms, scored from 0 (never) to 5 (very strongly). Pain course pattern, scored from –1 to 2 depending on pattern choice, and radiating pain (yes = 2, no = 0) are also assessed. The total PDQ score, which ranges from –1 to 38, is used to assess the probability of an existing neuropathic pain element, where scores ≤12 suggest a neuropathic component is unlikely (<15%), whereas scores ≥19 indicate a high likelihood of a neuropathic pain component (>90%). Scores between 12 and 19 are inconclusive^28^.

#### Knee Injury and Osteoarthritis Outcome Score 12 (KOOS-12)

In addition to assessing aspects of pain intensity and quality, the impact of pain on an individual’s ability to function and experience life is a critical outcome in pain management^32^. Understanding how pain interferes with daily functioning may help inform treatment decisions, assess the impact of pain on quality of life, and identify individuals who are at risk of chronic pain and disability^33–35^. The KOOS-12 was included to capture knee pain, difficulty with functional daily activities, and quality of life in the knee arthroplasty cohort. The KOOS-12 assesses each domain using Likert responses ranging from 0 ‘none’ to 4 ‘extreme’^36^. The pain subscale queries knee pain experienced in the past week while walking on a flat surface, going up or down stairs, and sitting or lying down. The function, daily living subscale queries the degree of difficulty experienced in the past week while rising from bed, standing, getting in/out of a car, and twisting/pivoting on the injured knee. The quality of life subscale measured an individual’s experience of knee pain-related quality of life. The subscale scores were transformed to a 0-100 scale where 0 represents no pain or dysfunction and 100 represents extreme pain or dysfunction.

#### Danish Thoracic Surgery Questionnaire

Individuals with moderate to severe postsurgical thoracic pain report significant limitations with daily activites^37–39^. The modified Danish Thoracic Surgery Questionnaire (DTSQ) was incorporated as an analogue to the KOOS-12 with a focus on capturing aspects of pain-related impairment and quality of life in the thoracic surgery cohort. The modified DTSQ is a 17-item scale which assesses functional impairment following thoracic surgery with item responses from 0 (“pain impairs me not at all”) to 4 (“I never do this activity due to pain”)^37,40^. A cumulative pain impairment score (0-68) was generated as the sum of the 17 responses.

### Evoked Pain Assessments

The peripheral and central nervous systems play an integral role in pain perception. Movement evoked pain (MEP) and QST may help characterize pain mechanisms, identify persons who are susceptible to the development of chronic pain, and inform targeted interventions. In particular, QST provides an avenue to indirectly examine somatosensory function. Evoked pain assessments performed pre-operatively and at the 3-month follow-up visit, generally performed in the following order: movement pain tasks, shoulder pressure pain thresholds (PPTs), dynamic mechanical allodynia (DMA, thoracic cohort only), PPTs at index site, mechanical temporal summation (MTS) at shoulder and index sites, and conditioned pain modulation (CPM) (see **Supplemental Materials, Figure S1**). The cuff pain assessments occurred during the imaging portion of the study. All pain ratings collected during evoked pain assessments relied on the 0 – 10 verbal NRS, with options for half numbers to increase precision. Anchors were defined as 0 = no pain, and 10 = worst pain imaginable. An additional movement pain assessemt occurred at the 6-week assessment in the thoracic cohort only. See **Table 2** for a list of evoked pain metrics and their corresponding abbreviations, brief summaries and dataset variables available to represent or generate each assessment.

**Table 2.** Evoked-Pain Measure Variables - List of Abbreviations.

| Measure | Abbreviation | Full Name | Dataset Variables Used |
| --- | --- | --- | --- |
| **QST: Pressure Pain Threshold (PPT)** | PPT Thor | Mean PPT Thoracic (Index for thoracic cohort) | primary_ppt_thor |
|  | PPT Knee | Mean PPT Knee (index for knee cohort) | primary_ppt_tka |
|  | PPT Shld | Mean PPT Shoulder (remote site) | secondary_ppt |
|  | MTS Thor Diff | MTS Thoracic Pain Rating Difference [(max – initial), index] | primary_ts_index_thor |
|  | MTS Thor Ratio | MTS Thoracic Pain Rating Ratio Change [(max +1)/(initial+1), index] | secondary_ts_index_thor |
| **QST: Mechanical Temporal Summation (MTS)** | MTS Knee Diff | MTS Knee Pain Rating Difference [(max-initial), index] | primary_ts_index_tka |
|  | MTS Knee Ratio | MTS Knee Pain Rating Ratio Change [(max +1)/(initial+1), index] | secondary_ts_index_tka |
|  | MTS Shld Diff | MTS Shoulder Pain Rating Difference [(max-initial), index] | primary_ts_remote |
|  | MTS Shld Ratio | MTS Shoulder Pain Rating Ratio Change [(max +1)/(initial+1), index] | secondary_ts_remote |
| **QST: Conditioned Pain Modulation (CPM)** | CPM Ratio | CPM PPT Ratio [(PPTpre - PPTpost)/PPT pre] | primary_cpm |
|  | CPM Diff | CPM PPT Difference (PPTpre - PPTpost) | secondary_cpm |
|  | CPM Peak Hand Pain | CPM Peak Hand Pain | peak_hand_pain |
|  | P4 Cuff Pain Begin | Individualized Cuff (target 3.5 - 4/10) Pain first 2 min of cuff | fmricuffpainbegin |
|  | 120 Cuff Pain Begin | Standardized 120mmHg Cuff Pain first 2 min of cuff | Fmricuffpaincpbegin |
| **QST/fMRI: Cuff Pressure Pain** | P4 Cuff TS Diff | Individualized Cuff TS (final or max pain – beginning pain) | TSp4cuff |
|  | 120 Cuff TS Diff | Standardized Cuff TS (final or max - beginning) | TS120cuff |
|  | P4 Cuff Pressure | Individualized Cuff Pressure to achieve 3.5 - 4/10 pain (mm Hg) | cuff_pain4pressure |
| **QST: Dynamic Mechanical Allodynia (DMA) †** | DMA Cont | DMA Mean Pain Rating for Contralateral Thoracic Site | dmacont |
|  | DMA Thor | DMA Mean Pain Rating for Ipsilateral Thoracic Site (Surgical Side) | Dmaindx |
| **5 Times Sit-to-Stand (5TSTS) \*** | 5TSTS Pain Max | 5TSTS Maximum Pain Rating | tstspostpainscl |
|  | 5TSTS MEP | 5TSTS Movement Evoked Pain (Max – Resting ) | mep_5tsts |
| **10 Meter Walk Test (10MWT) \*** | 10MWT Pain Max | 10MWT Maximum Pain Rating | walk10finalpainscl |
|  | 10MWT MEP | 10MWT Movement Evoked Pain (Max – Resting) | mep_walk |
| **Deep Breathing †** | Breath Pain Max | Deep Breathing Maximum Pain Rating | ftdbcdeepbrthfinalscl |
|  | Breath MEP | Deep Breathing Movement Evoked Pain (Max – Resting) | mep_breath |
| **Coughing †** | Cough Pain Max | Coughing Maximum Pain Rating | ftdbccoughfinalscl |
|  | Cough MEP | Coughing Movement Evoked Pain (Max – Resting) | mep_cough |

†Thoracic cohort only. \*Knee cohort only.
QST = Quantitative sensory testing; fMRI = functional magnetic resonance imaging

#### Movement Evoked Pain

Movement-evoked pain are increasingly recognized as constructs that are distinct from pain at rest and are associated with poorer post-operative pain and functional outcomes ^41^. MEP was evaluated in two ways: the primary metric was the change score, defined as the difference between initial pain and maximal pain with activity which represents pain “evoked” by the movement. Evidence also supports that peak pain with movement is an important construct^42–44^, thus maximum pain during each task was chosen as a secondary outcome – referred to as movement pain. Movement tasks were selected for each cohort for their clinical relevance and ease of administration (see below for details). Knee or chest pain was assessed prior to each task (0 – 10 NRS using half or whole numbers) as appropriate, with the maximum pain experienced during the activity assessed upon completion.

For the knee cohort, movement pain and MEP were assessed using two standard performance tasks: the Five-Times Sit-to-Stand (5TSTS) test^45,46^ and the 10-meter Walk Test (10MWT)^47,48^. Each activity was performed following standard protocols, with pain ratings collected both before and after each task^45–50^. In addition to MEP metrics, the performance outcomes for each activity were recorded (i.e., time to completion).

For the thoracic cohort, movement pain and MEP were assessed using two respiratory maneuvers: deep breathing and coughing, reflecting common postoperative care protocols^51^. Pain was assessed prior to and following each task. Participants first completed three diaphragmatic deep breaths, inhaling through the nose and exhaling slowly through the mouth after a hold of 3 seconds. The coughing task involved asking participants to breathe in deeply, and perform 2-3 hard forceful coughs. Only pain metrics were assessed for these tasks, no other performance metrics (e.g., volumetric measures) were evaluated.

#### Pressure Pain Thresholds

Pressure pain sensitivity is related to clinical pain severity in individuals with knee osteoarthritis^52,53^ and post-operative pain following knee arthoplasty^54–56^ and thoracic surgery^57–59^. These studies suggest that individuals with lower PPTs exhibit higher pressure pain sensitivity and are at higher risk for experiencing severe pain post-operatively and transitioning to chronic pain. PPTs were performed locally at the index surgical site (knee or chest) and at a standard remote site (middledeltoid of the shoulder contralateral to the surgical site). The index knee site was performed at the mid-portion of the medial or lateral joint line, corresponding to the participant’s reported location of worst pain. If pain was reported as equal between medial and lateral compartments or as anterior knee pain, the default PPT location was medial joint line. For the thoracic cohort, the index PPT site was located between the 5^th^ and 6^th^ intercostal space along the mid axillary line.

PPTs were assessed using a manual pressure algometer (Wagner Pain Test FPX25, Wagner Instruments, USA), with a 1-cm^2^ rubber tip applied perpendicularly to the skin at a rate of 0.5 kgf/s until faint pain (i.e approximately 1 out of 10) was noted. Three assessments were performed at each site with a brief rest interval (∼10 sec) and slight overlap between assessment sites. If a PPT assessment yielded an unusual value or appeared questionable (e.g., due to movement or algometer slippage), the test was repeated, and the final three pressure ratings were averaged to obtain a PPT value for each site.

#### Mechanical Temporal Summation

Temporal summation of pain provides an indication of the heightened perception of pain with repeated painful stimuli and is an indirect measure of central pain facilitation^60^. Temporal summation, when measured before surgery, is a strong predictor of the intensity of both acute and chronic pain following surgery, notably in knee arthroplasty^52,61,62^ and thoracic surgery^59,63^. MTS of pain was assessed at the surgical (index) and remote (shoulder) sites using a punctate stimulus (Neuropen®, Owen Mumford, United Kingdom). The Neuropen was applied approximately 1 cm away from the PPT sites to prevent any overlap (i.e., superior to the PPT site for the remote site, inferior for the index site). The Neuropen was applied perpendicularly to the skin, with the probe depressed per manufacturer’s instructions to the marked level to apply standardized pressure (40 g). The remote site was tested first by applying a single poke and asking the participant to rate the pain using a 0 – 10 verbal pain NRS. The Neuropen was subsequently applied 10 times at a rate of 1 Hz within a 1 cm radius, followed by a maximum pain rating. The protocol was repeated three times at each site. After the third (final) trial for each site, after-sensations of pain (0-10 NRS) were recorded at 15 and 30 sec. The primary outcome was calculated as the mean of the three MTS difference scores (maximal pain evoked during the 10 pokes minus pain evoked with a single poke) at the surgical index site. The mean of the three MTS difference scores at the remote (shoulder) site was a secondary outcome. Additionally, a MTS ratio score was calculated as ([max pain during 10 pokes +1]/ [pain with a single poke +1]).

#### Conditioned Pain Modulation

Conditioned pain modulation (CPM) provides an indirect measure of central pain inhibition^64–66^. Attenuated CPM prior to surgery has been associated with increased post-operative pain across a number of conditions, including knee replacement^67,68^ surgery and thoracotomy^69,70^. The CPM paradigm used in this study assessed the change in PPT at the remote (shoulder) site after submersion of the hand/wrist ipsilateral to the index site for up to 1 min in a circulating cold-water bath maintained at 10° +/-1°C. Each site had different water bath equipment, but all met these study protocol requirements. PPTs previously assessed at the beginning of the QST session were used as the baseline (pre-water submersion) values. Cold pain ratings were assessed at 30 and 60 sec, or at the time of hand withdrawal if less than 60 sec; with the maximum pain extracted for analysis. Following hand withdrawal, PPTs were immediately assessed three times at the remote (shoulder) site as previously described. If participants were unable to complete the full 1-minute, the duration were documented. The primary CPM outcome was computed as the percent change (see Eq 1 below); the secondary CPM outcome was the difference score (pre-PPT - post-PPT).

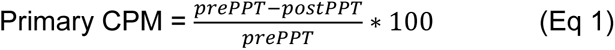

#### Dynamic Mechanical Allodynia

Allodynia has been reported as a symptom of post-thoracotomy pain syndrome^71–73^ and accordingly dynamic mechanical allodynia was assessed in the thoracic cohort only. While allodynia is commonly associated with neuropathic pain, direct nerve injury is not required to produce central sensitization and its associated pain with non-painful stimuli such as light touch and movement^71,74^. Allodynia was assessed at a standardized location on the lateral thorax at the midaxillary line between the 5^th^ and 6^th^ intercostal spaces, ipsilateral and contralateral to the surgical site at baseline and 3-month visits. Additionally at the 3-month post-surgical visit, a second DMA site was marked at the most painful or sensitive area on their chest wall unless the identified site overlapped with the standardized index site (< 4 cm). If the participants denied any pain or sensitivity, the second DMA site was marked 2 cm inferior to the midpoint of the surgical scar (most anterior scar if more than one). DMA was assessed by lightly stroking the identified sites five times with a soft-bristle brush (flat synthetic filament brush, width 16 mm and length 19 mm) in a posterior to anterior direction. Each stroke was performed with the bristles bent to approximately 45 degrees, with a 4-cm stroke length over 1 sec. Pain ratings (0-10 NRS) were elicited after each brush stroke. DMA was calculated as the mean of the five pain ratings at each site, where a mean score >0 would indicate allodynia.

#### Cuff Pressure Pain and Temporal Summation

Similar to PPT, cuff pressure pain has been associated with postoperative pain^75^. As part of the A2CPS functional imaging protocol, two continuous 6-min cuff pressure conditions were assessed. The first was a subject-specific pressure level calibrated to evoke a pain intensity rating of approximately 4/10 (P4 cuff), and the second was a pre-selected standard pressure (120 mmHg) for all participants (120 cuff). The cuff was applied to the calf of the non-dominant leg (or dominant leg if contraindications existed) for the thoracic surgery cohort and to the leg contralateral to the surgical knee in the knee arthroplasty cohort. Immediately following both 6-minute imaging sequences, participants were asked to recall cuff-evoked pain intensity (0-10 NRS) at the calf for the first two minutes, middle two minutes, and last two minutes. Pain sensitivity from the cuff was assessed in several ways: 1) pressure needed to elicit approximately 4/10 pain (P4 cuff); 2) initial cuff pain ratings for the 120 mmHg condition (120 cuff); and 3) MTS for both conditions computed as maximum pain (during the middle 2 min and last 2 min of inflation) minus pain from first 2 min of inflation.

### A2CPS Data Quality Assurance and Quality Control

#### Research Assistant Training & Certification

To ensure consistent data collection for pain-related assessments, all study research assistants were required to complete comprehensive training, certification, and re-certification in conducting and recording pain assessments, performing inter-rater reliability evaluations, and reviewed for data completeness prior to site activation and every six months thereafter. More details on training and certification will be presented in a separate manuscript.

#### Live Data Monitoring with Site Feedback

The Data Integration and Resource Center continually monitored data completion for all pain assessments throughout the course of the study. This included monitoring data completeness across pain measures at all sites and notified research assistants at clinical sites of missing data. All sites received weekly reports of missing data, and concerning patterns in missingness were reviewed with research staff. This process helped ensure the collection of high-quality, consistent pain-related data across the study.

#### Missing Data Management

Both raw and imputed data are available within A2CPS data releases. In cases of missing data, each measure is handled on a case-by-case basis using available literature or user guides. Documentation describing score calculation and missing data imputation for each pain and pain sensitivity measure is available with the data download.

### Statistical Analyses

Descriptive statistics were performed on all pain and pain sensitivity variables and are presented as mean [standard deviation] or frequencies, n(%), as appropriate. Baseline differences between cohorts (knee vs. thoracic) were examined for measures common to both cohorts using Welch’s two-sample t-tests for continuous variables and Pearson’s chi-squared tests or Fisher’s exact test for categorical variables, as appropriate. Sex differences (male vs. female) were examined within each cohort as a secondary descriptive analysis using the same approach. Because these analyses were primarily descriptive, p-values should be interpreted cautiously with respect to multiple comparisons.

Pairwise associations among pain and pain sensitivity measures were summarized using correlation matrices and visualized with heat maps. Pearson correlation coefficients (r) were calculated separately for the combined cohort using measures common to both surgical groups and within each cohort using all available cohort-specific measures. Correlation analyses were restricted to participants with non-missing data for the variables included in each pairwise comparison. Variables were reviewed for distributional characteristics, floor effects, and near-zero variance prior to multivariable analyses.

Exploratory factor analysis (EFA) was used to characterize the latent structure of baseline pain and pain sensitivity measures. Prior to extraction, suitability for factor analysis was evaluated using the Kaiser–Meyer–Olkin (KMO) measure of sampling adequacy, Bartlett’s test of sphericity, and consideration of missingness and communality. As part of a prespecified variable-screening process, measures with substantial missingness (> 40%), sampling inadequacy (item level MSA < 0.50, near zero variance, or low communality (h^2^ < 0.20) were excluded before factor extraction. EFA was performed in the combined cohort using measures shared across cohorts and separately within each cohort using cohort-specific measures. Factors were extracted using principal axis factoring and rotated using oblimin allowing factors to correlate and facilitate interpretability. We used parallel analysis, visual inspection of the parallel-analysis scree plot, and the interpretability and theoretical coherence of the resulting solutions to determine the number of factors. For parallel analysis we used principal-axis (common-factor) eigenvalues, in which the observed factor eigenvalues were compared against those from 100 randomly simulated and resampled datasets; a factor was retained when its observed eigenvalue exceeded the corresponding parallel-analysis eigenvalue. In some cases, the model suggested by parallel analysis failed to converge or yielded an ultra-Heywood case (communality exceeded 1.0). To offset this, we retained a smaller number of factors that produced a stable, interpretable solution.

Correlation Network Graphs of pain variables within and across each latent domain were generated using the visNetwork package in R^76^. The Nodes represented individual measures, colored according to their factor assignment. The edges between the nodes represented bivariate Pearson correlations that exceeded a minimum threshold of |r| ≥ 0.10 for the combined cohort and |r| ≥ 0.20 for the cohort-specific analyses. Edge width was determined by the magnitude of the correlation coefficient, and positive vs. negative associations were differentiated by the edge color. Because some pain measures were either sparse, or did not follow a normal distribution, the networks were used as an exploratory tool only in conjunction with the factor analysis.

## Results

Baseline data from a total of 1,401 participants were included in these analyses (data release 2.1.0), of which 1,043 were in the knee cohort and 358 in the thoracic cohort (**Table 3**). The proportion of females was higher in both groups (Knee: 63% female; Thoracic: 56% female). Individuals in the knee cohort were older on average (mean ± SD: 65.2 ± 8.5 years) compared to the thoracic cohort (59.8 ± 13.0 years). We highlight several pain and pain sensitivity baseline metrics below, with more details provided in the respective tables.

**Table 3.**
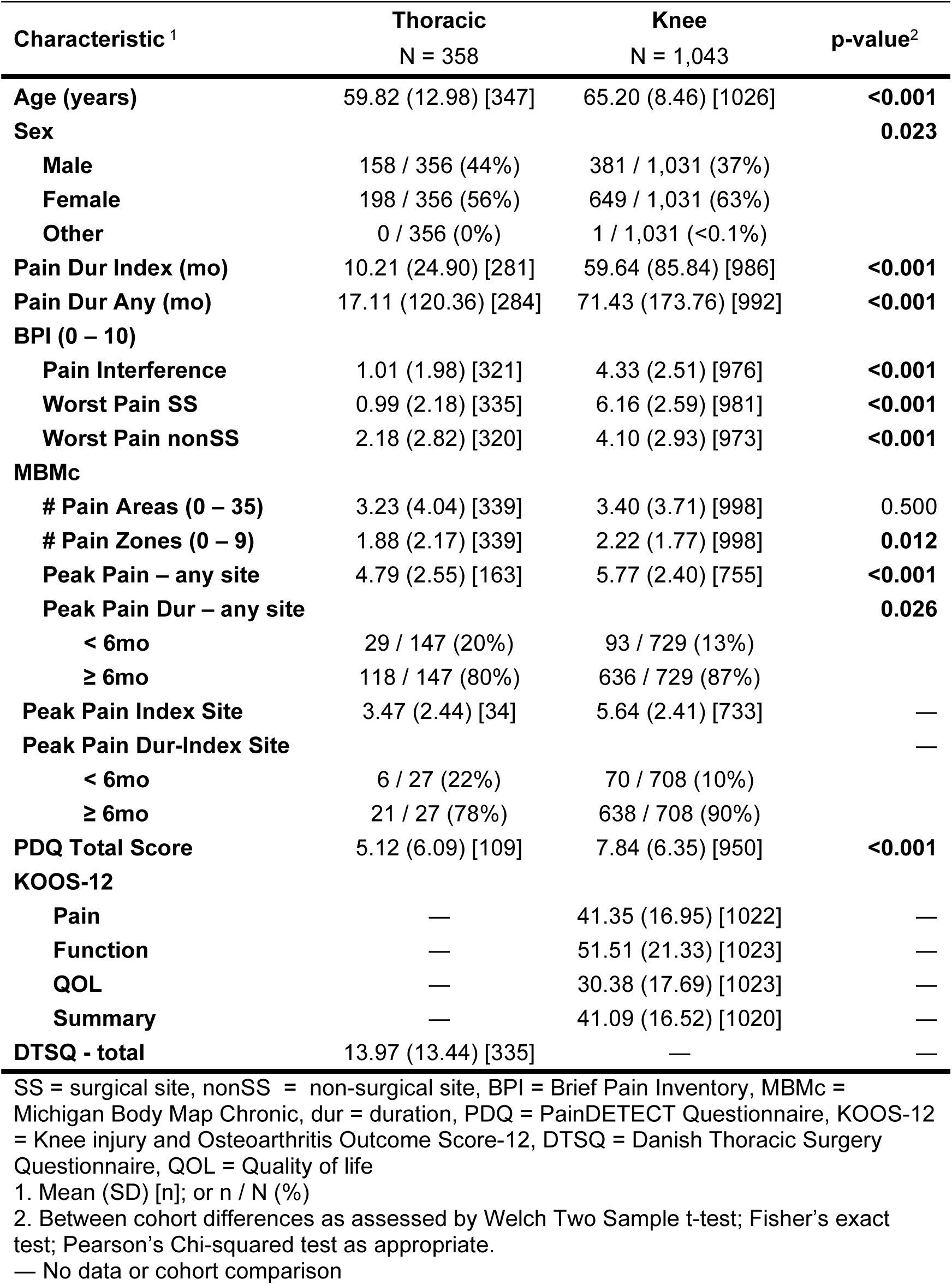
Summary Participant Characteristics & Self-Report Pain Measures by Cohort.

| Characteristic <sup>1</sup> | Thoracic<br>N = 358 | Knee<br>N = 1,043 | p-value<sup>2</sup> |
| --- | --- | --- | --- |
| **Age (years)** | 59.82 (12.98) [347] | 65.20 (8.46) [1026] | **<0.001** |
| **Sex** |  |  | **0.023** |
| **Male** | 158 / 356 (44%) | 381 / 1,031 (37%) |  |
| **Female** | 198 / 356 (56%) | 649 / 1,031 (63%) |  |
| **Other** | 0 / 356 (0%) | 1 / 1,031 (<0.1%) |  |
| **Pain Dur Index (mo)** | 10.21 (24.90) [281] | 59.64 (85.84) [986] | **<0.001** |
| **Pain Dur Any (mo)** | 17.11 (120.36) [284] | 71.43 (173.76) [992] | **<0.001** |
| **BPI (0 – 10)** |  |  |  |
| **Pain Interference** | 1.01 (1.98) [321] | 4.33 (2.51) [976] | **<0.001** |
| **Worst Pain SS** | 0.99 (2.18) [335] | 6.16 (2.59) [981] | **<0.001** |
| **Worst Pain nonSS** | 2.18 (2.82) [320] | 4.10 (2.93) [973] | **<0.001** |
| **MBMc** |  |  |  |
| **# Pain Areas (0 – 35)** | 3.23 (4.04) [339] | 3.40 (3.71) [998] | 0.500 |
| **# Pain Zones (0 – 9)** | 1.88 (2.17) [339] | 2.22 (1.77) [998] | **0.012** |
| **Peak Pain – any site** | 4.79 (2.55) [163] | 5.77 (2.40) [755] | **<0.001** |
| **Peak Pain Dur – any site** |  |  | **0.026** |
| < 6mo | 29 / 147 (20%) | 93 / 729 (13%) |  |
| ≥ 6mo | 118 / 147 (80%) | 636 / 729 (87%) |  |
| **Peak Pain Index Site** | 3.47 (2.44) [34] | 5.64 (2.41) [733] | — |
| **Peak Pain Dur-Index Site** |  |  | — |
| < 6mo | 6 / 27 (22%) | 70 / 708 (10%) |  |
| ≥ 6mo | 21 / 27 (78%) | 638 / 708 (90%) |  |
| **PDQ Total Score** | 5.12 (6.09) [109] | 7.84 (6.35) [950] | **<0.001** |
| **KOOS-12** |  |  |  |
| **Pain** | — | 41.35 (16.95) [1022] | — |
| **Function** | — | 51.51 (21.33) [1023] | — |
| **QOL** | — | 30.38 (17.69) [1023] | — |
| **Summary** | — | 41.09 (16.52) [1020] | — |
| **DTSQ - total** | 13.97 (13.44) [335] | — | — |

SS = surgical site, nonSS = non-surgical site, BPI = Brief Pain Inventory, MBMc = Michigan Body Map Chronic, dur = duration, PDQ = PainDETECT Questionnaire, KOOS-12 = Knee injury and Osteoarthritis Outcome Score-12, DTSQ = Danish Thoracic Surgery Questionnaire, QOL = Quality of life
1. Mean (SD) [n]; or n / N (%)
2. Between cohort differences as assessed by Welch Two Sample t-test; Fisher's exact test; Pearson's Chi-squared test as appropriate.
— No data or cohort comparison

### Self-Report Pain Surveys

At baseline, individuals in the knee cohort reported substantially higher pain than those in the thoracic cohort for worst pain at the planned surgical site (6.2 ± 2.6 vs.1.0 ± 2.2) and non-surgical sites (4.1 ± 2.9 vs. 2.2 ± 2.8). They also reported higher pain interference (4.3 ± 2.5) compared with the thoracic cohort (1.0 ± 2.0). See **Table 3** for more details on baseline self-reported pain metrics in both cohorts.

Despite these differences in pain severity and interference, the number of chronic pain areas (out of 35 possible areas) endorsed on the MBM was similar between groups (p = 0.5). However, when reduced to 9 zones, the knee cohort reported slightly more widespread pain than the thoracic cohort: 2.2 ± 1.8 vs. 1.9 ± 2.2 (p = 0.012). Participants in both cohorts reported moderate to severe peak pain anywhere in their body, but was significantly higher in the knee than the thoracic cohort (5.8 ± 2.4 vs. 4.8 ± 2.6, p < 0.001). Further, most participants reported this site of peak pain was present for more than 6 months (87% and 80% for knee and thoracic cohorts, respectively). Additionally, the knee cohort endorsed greater neuropathic symptoms (7.5 ± 6.4) relative to the thoracic cohort (5.1 ± 6.1) on the PainDETECT (P < 0.001).

For condition-specific measures, the knee cohort demonstrated a mean KOOS-12 summary score of 41.1 ± 16.5, indicative of substantial knee-related symptoms and functional impairment pre-surgery, whereas the thoracic cohort had a mean DTSQ summary score of 14.0 ± 13.4, suggesting mild functional impairment pre-surgery.

### Evoked-Pain Measures

#### Movement-Evoked Pain

Participants in the knee cohort reported mild to moderate pain during the function tests (3.7 ± 2.6 and 3.1 ± 2.6, for 5TSTS and 10MWT, respectively, **Table 4**). In contrast, minimal pain was noted in the thoracic cohort, 0.4 ± 1.1 and 0.6 ± 1.4 during deep breathing and coughing, respectively. For the MEP difference score, knee participants reported increased pain during both functional tasks:1.9 ± 2.0 for the 5TST and 0.7 ± 1.3 for the 10MWT over baseline pain. In contrast, thoracic participants reported minimal to no pain increase during deep breathing (0.05 ± 0.6) or coughing (0.2 ± 0.9) over baseline pain.

**Table 4.** Pain with Activity by Cohort Measured with a 0-10 Numerical Rating Scale.

|  | **Thoracic** |  | **Knee** |
| --- | --- | --- | --- |
|  | **N = 358<sup>1</sup>** |  | **N = 1,043<sup>1</sup>** |
| **Max Pain with Movement:** |  |  |  |
| **Deep Breathing** | 0.38 (1.10) [356] | **5TSTS** | 3.68 (2.62) [992] |
| **Coughing** | 0.56 (1.37) [356] | **10MWT** | 3.13 (2.56) [1018] |
| **Movement-Evoked Pain** |  |  |  |
| **Deep Breathing** | 0.05 (0.58) [356] | **5TSTS** | 1.85 (1.98) [991] |
| **Coughing** | 0.23 (0.88) [356] | **10MWT** | 0.67 (1.28) [1015] |

5TSTS = Five times sit to stand test; 10MWT = 10-meter walk test; Pain
Max = Peak pain with movement; MEP = movement-evoked pain (difference between max and initial resting pain).
1. Total cohort sample size; available data for each characteristic provided as [n].

#### Pressure Pain Thresholds

At the respective index sites, mean PPTs were 2.8 ± 1.7 kgf/cm^2^ for the knee cohort (knee site) and 2.3 ± 1.4 kgf/cm^2^ for the thoracic cohort (chest site). PPTs did not differ at the common remote shoulder site, with mean and SD values of 3.2 ± 1.8 kgf/cm^2^ for the knee and 3.3 ±1.8 kgf/cm^2^ for the thoracic cohort. (p = 0.3, see **Table 5**)

**Table 5.** Quantitative Sensory Testing Measures by Cohort.

| Characteristic | Thoracic<br>N = 358<sup>1</sup> | Knee<br>N = 1,043<sup>1</sup> | p-value<sup>2</sup> |
| --- | --- | --- | --- |
| **Pressure Pain Threshold (PPT) (kgf)** |  |  |  |
| Index site | 2.32 (1.39) [348] | 2.79 (1.66) [1018] | — |
| Shoulder | 3.34 (1.84) [348] | 3.23 (1.76)[1020] | 0.3 |
| **Mechanical Temporal Summation (MTS)** |  |  |  |
| Index site - Difference | 1.67 (1.65) [345] | 2.36 (1.80) [987] | — |
| Index site - Ratio | 2.17 (1.26) [345] | 2.29 (1.19) [987] | — |
| Shoulder - Difference | 1.11 (1.17) [345] | 1.63 (1.50) [1006] | **<0.001** |
| Shoulder - Ratio | 1.84 (0.88) [345] | 2.12 (1.15) [1006] | **<0.001** |
| **Conditioned Pain Modulation (CPM)** |  |  |  |
| Ratio (%) | -11.85 (35.54) [334] | -7.21 (34.34) [987] | **0.038** |
| Difference (kgf) | -0.28 (0.95) [334] | -0.11 (0.94) [987] | **0.004** |
| Peak Hand Pain (0-10) | 6.46 (2.81) [334] | 6.00 (2.98) [1007] | **0.012** |
| **Cuff Pressure Pain** |  |  |  |
| P4 Cuff Pain Begin (0-10) | 3.19 (1.92) [170] | 3.32 (2.06) [557] | 0.5 |
| 120 Cuff Pain Begin (0-10) | 2.62 (2.13) [157] | 2.95 (2.15) [479] | 0.1 |
| P4 Cuff TS Difference | 1.64 (2.14) [169] | 1.72 (2.16) [554] | 0.7 |
| 120 Cuff TS Difference | 0.89 (1.98) [157] | 1.28 (1.98) [476] | **0.032** |
| P4 Cuff Pressure (mmHg) | 164.24 (63.88) [311] | 137.01 (55.00) [732] | **<0.001** |
| **Dynamic Mechanical Allodynia (DMA)** |  |  |  |
| Index | 0.01 (0.08) [348] | — | — |
| Contralateral Chest | 0.00 (0.06) [347] | — | — |

P4 Cuff = individualized pain 4/10 cuff condition, 120 Cuff = standardized 120mmHg cuff condition
1. Total cohort sample size; available data for each characteristic provided as [n].
2. Differences between cohorts assessed by Welch Two Sample t-test as appropriate.
— No data or no cohort comparison performed.

#### Mechanical Temporal Summation

The primary MTS outcomes (increase in pain following 10 stimuli) at the respective index sites were 2.4 ± 1.8 for the knee cohort and 1.7 ± 1.7 for the thoracic cohort. At the common shoulder site, MTS was slightly, but significantly, greater in the knee cohort than the thoracic cohort, for both the difference scores (1.6 ± 1.5 versus 1.1 ± 1.2) and the wind-up ratios (2.1 ± 1.2 versus 1.8 ± 0.9), respectively.

#### Conditioned Pain Modulation

Both cohorts experienced a slight increase in pain thresholds (i.e., pain inhibition) following cold water hand immersion. However, CPM inhibition was greater in the thoracic cohort than the knee cohort using both primary (ratio percentage, p = 0.038) and secondary (difference score, p = 0.004) CPM metrics (ratio: −11.9 ± 35.5% versus −7.2 ± 34.3%; and difference: −0.3 ± 1.0 kgf/cm^2^ versus −0.1 ± 0.9 kgf/cm^2^ for thoracic and knee cohorts, respectively).

#### Cuff Pain and Temporal Summation

The individualized cuff pressure calibrated to evoke moderate pain (target 3.5 - 4.0/10 pain) was greater in the thoracic group (164.24 ± 63.88 mmHg) compared to the knee cohort (137.01 ± 55.00 mmHg). Both cohorts reported achieving similar cuff pain during the first two minutes of inflation during the fMRI procedure (3.3 ± 2.1 and 3.2 ± 1.9 for knee and thoracic cohorts, respectively, p = 0.5). Both cohorts responded similarly to this pressure over the 6 min, with pain increases (i.e., temporal summation difference scores) of 1.7 ± 2.2 for knee and 1.6 ± 2.1 for thoracic cohorts. During the standard pressure (120 mmHg) cuff protocol, however, the observed MTS of cuff pain was greater in the knee cohort (1.3 ± 2.0) than in the thoracic cohort (0.9 ± 2.0; p = 0.032). See **Table 5** for more details.

### Sex Differences

In the knee cohort, female participants reported higher BPI pain intensity (p < 0.001), greater BPI pain interference (p < 0.001), more widespread pain (p = 0.013), and more neuropathic pain symptoms (p = 0.036) than male participants (see **Supplemental Materials, Table S1 to S3**). Similarly, males had consistently higher KOOS scores, reflecting less pain and pain interference, than females (p < 0.001). In the thoracic cohort, females reported more widespread pain (p = 0.021), higher pain intensity at sites outside the chest wall (p < 0.001), greater impairment on the DTSQ (p < 0.001), and more neuropathic pain symptoms (p = 0.029) than males, but no significant sex differences in pain interference or thoracic site pain (both p ≥ 0.13, see **Supplemental Materials, Table S4 to S6**).

### Correlation Among Pain and Pain Sensitivity Measures

Correlations among common clinical pain and evoked pain sensitivity measures for the entire cohort are presented in **Figure 1**. The figure illustrates both positive and negative associations, reflecting that some metrics increase while others decrease with higher levels of pain and pain sensitivity. The heat map highlights clusters of measures that tend to co-vary—for example, BPI, MBMc, and PDQ metrics group together and show similar relationships, whereas the cuff-based pain metrics form a separate cluster. In contrast, several measures, such as CPM, show minimal or no correlation with the other variables.

**Figure 1.**
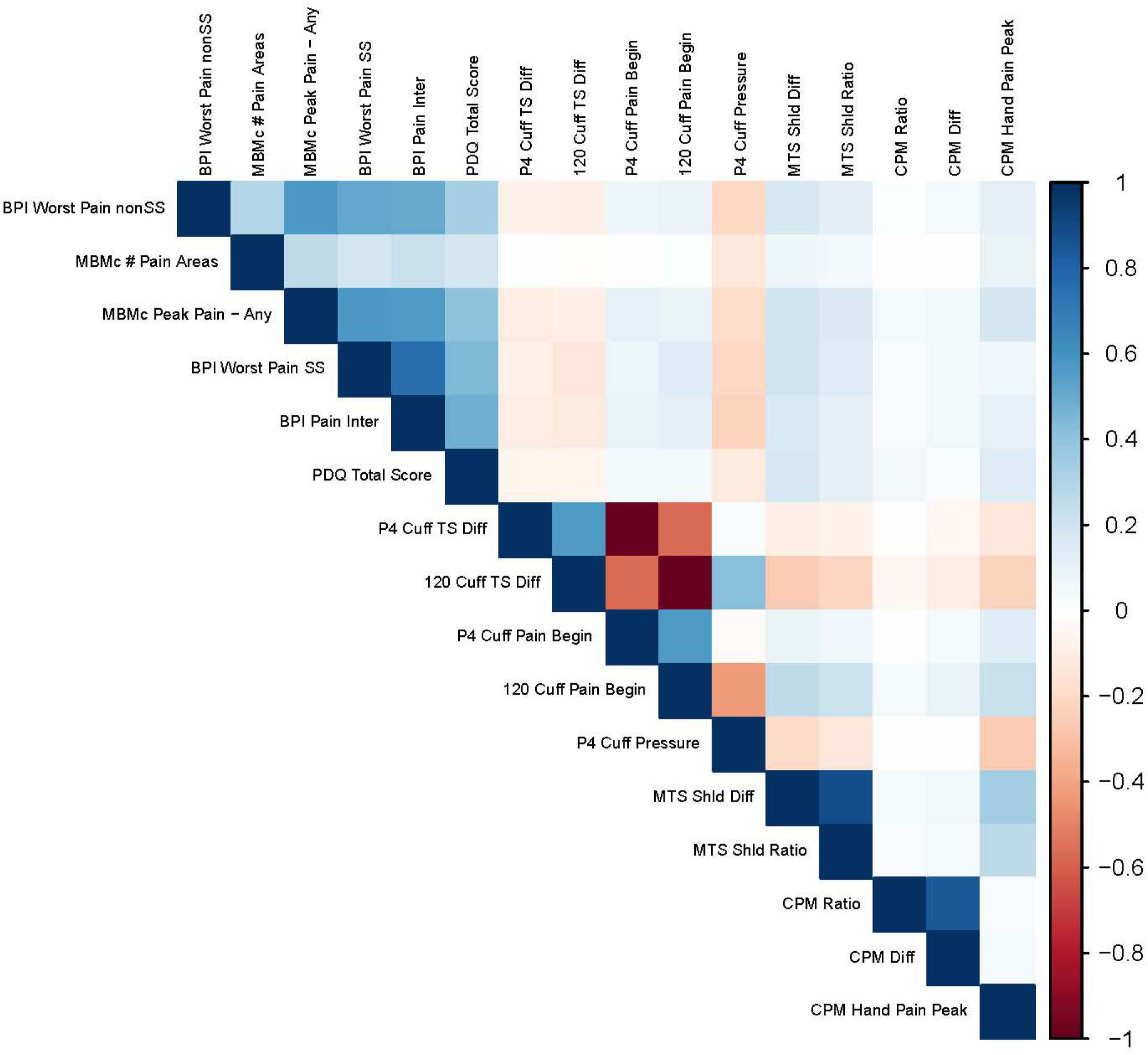
Pearson correlation heat map for including both cohorts, showing which pain and pain sensitivity metrics most strongly correlate positively (dark blue) or negatively (dark red), with minimal correlation between metrics represented as white. BPI = Brief Pain Inventory, nonSS = non-surgical site, MBMc = Michigan Body Map chronic, SS = surgical site, Inter = interference, PDQ = PainDETECT Questionnaire, P4 Cuff = individualized pain 4/10 cuff condition, 120 Cuff = standardized 120mmHg cuff condition, TS = temporal summation, Diff = difference score, MTS = mechanical temporal summation, Shld = shoulder, CPM = conditioned pain modulation.

Cohort-specific correlation heat maps are presented in **Figure 2**. Weak to moderate correlations were observed within each cohort, with generally greater associations and variable clusters observed in the knee than the thoracic cohorts using these assessments. Clusters of pain and pain sensitivity variables typically grouped by their underlying assessment method, but not always. For example, in the knee cohort, movement pain (i.e., maximum pain during movement) more closely associated with BPI and MBM pain intensity than with the MEP difference score from the same test. Correlation figures further separated by sex are available in Supplemental Materials, Figures S2 to S3 for the knee and thoracic cohorts.

**Figure 2.**
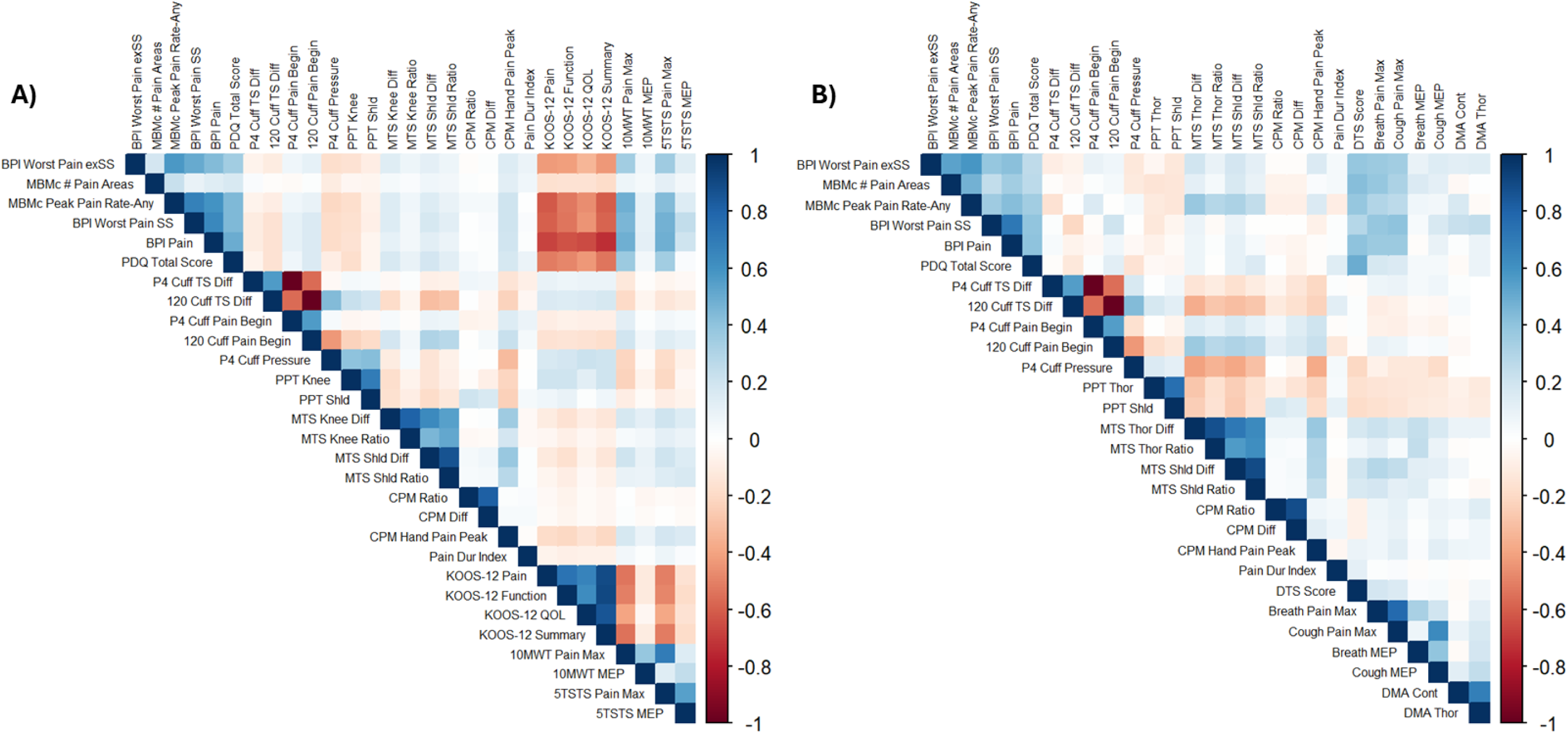
Pearson correlation heat map for A) knee and B) thoracic cohorts, showing which pain and pain sensitivity metrics most strongly correlate positively (dark blue) or negatively (dark red), with minimal correlation between metrics represented as white. BPI = Brief Pain Inventory, exSS = excluding surgical site, MBMc = Michigan Body Map chronic, SS = surgical site, PDQ = PainDETECT Questionnaire, P4 Cuff = individualized pain 4/10 cuff condition, 120 Cuff = standardized 120mmHg cuff condition, TS = temporal summation, Diff = difference score, PPT = pressure pain threshold, MTS = mechanical temporal summation, Shld = shoulder, CPM = conditioned pain modulation, Dur = duration, KOOS-12 = Knee injury and Osteoarthritis Outcome Score-12, QOL = Quality of life, 10MWT = 10 meter walk test, MEP = movement-evoked pain, 5TSTS = Five Times Sit-to-Stand, Thor = thoracic, Cont = control site, DMA = dynamic mechanical allodynia.

### Exploratory Factor Analysis

The remaining variables that met completeness, sampling adequacy, and communality thresholds formed the final reduced, and stable set of 16 shared measures, which were entered into the EFA for combined knee and thoracic cohorts (**Supplemental Materials, Figure S4**). When analyzed individually, the knee and thoracic cohorts included 21 and 16 measures in their final reduced EFAs, respectively (see **Supplemental Materials, Figures S5 – S6**). For the combined cohort EFA, a clear five-factor solution emerged, with strong within⍰factor loadings and minimal cross-loadings (**Figure 3**). These latent factors are described briefly as follows:

**Figure 3.**
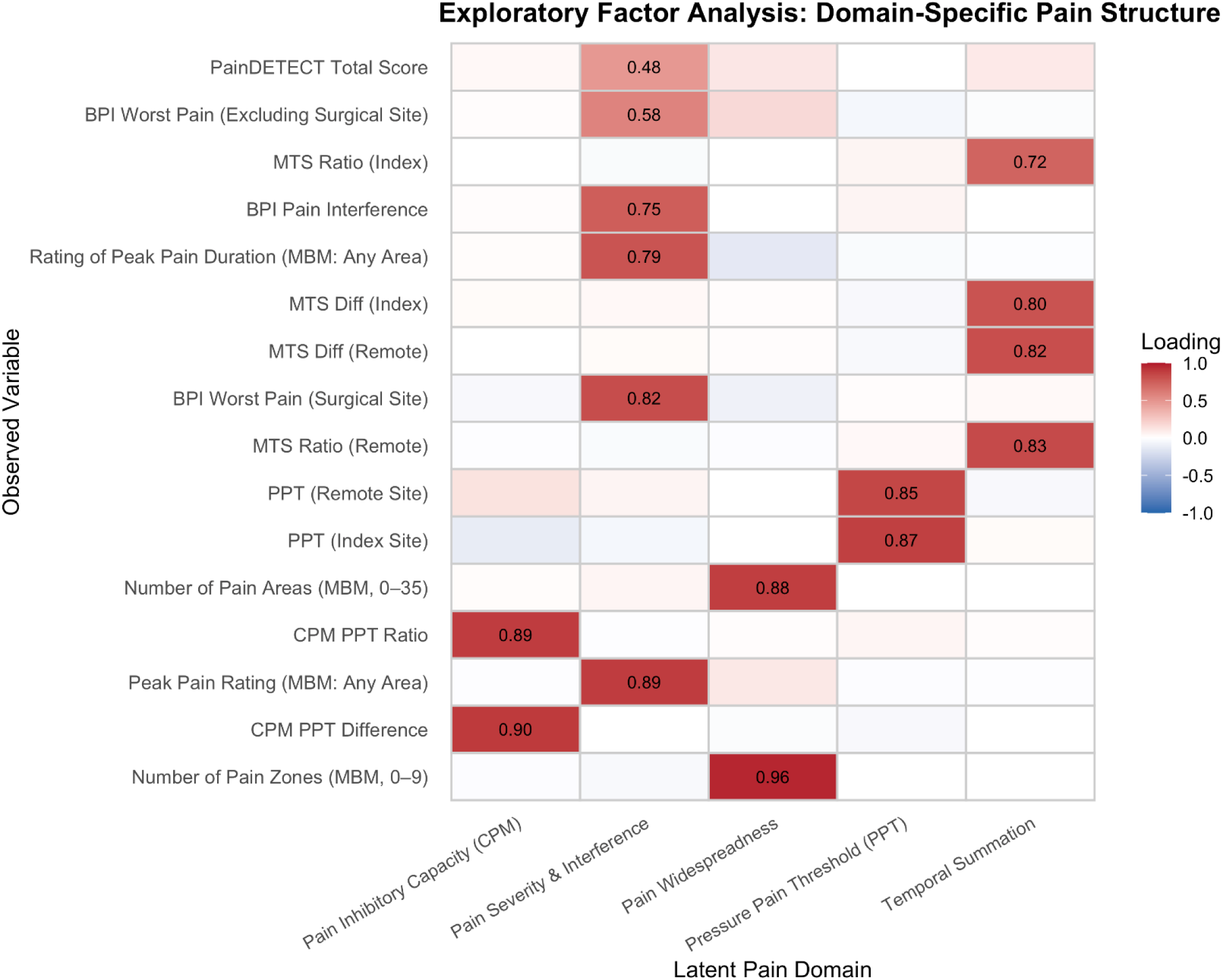
Summary of exploratory factor analysis using pain and pain sensitivity metrics similar across both cohorts indicating five latent pain domains. BPI = Brief Pain Inventory, MTS = mechanical temporal summation, MBM = Michigan Body Map, Diff = difference score, PPT = pressure pain thresholds, CPM = conditioned pain modulation.

*Factor 1: Pain Inhibitory Capacity.* CPM PPT Difference (0.90) and CPM PPT Ratio (0.89) loaded exclusively on this factor, representing endogenous inhibitory function.

*Factor 2: Pain Severity & Interference.* Self-reported pain measures—including BPI Pain Interference (0.75), BPI Worst Pain at surgical (0.82) and nonsurgical sites (0.58), peak MBM pain rating (0.89), rating of peak pain duration (0.79), and PainDETECT (0.48)—clustered together, reflecting a coherent pain-burden domain.

*Factor 3: Pain Widespreadness*. Spatial pain distribution formed a separate factor, dominated by number of pain areas (0.88) and number of pain zones (0.96).

*Factor 4: Pressure Pain Threshold*. PPT at both the index (0.87) and remote (0.85) sites loaded strongly, defining a mechanical pain⍰sensitivity domain even when index sites are distinct.

*Factor 5: Temporal Summation.* All MTS measures—ratio and difference at index and remote sites (0.72–0.83)—loaded together, indicating a unique excitatory pain-facilitation domain.

The cohort-specific EFA loadings are provided in **Figure 4A and B**. Across both cohorts, temporal summation, pressure pain threshold, pain inhibitory capacity, and pain widespreadness formed consistent latent domains, closely matching the structure identified in the full cohort. The key difference in the knee cohort, was the pain severity and interference factor split into separate factors, resulting in a 6-factor solution. Whilst the thoracic cohort, showed four common domains, accompanied by a distinct pain impact and movement-evoked pain (MEP) domain, yielding a 5-factor solution.

**Figure 4.**
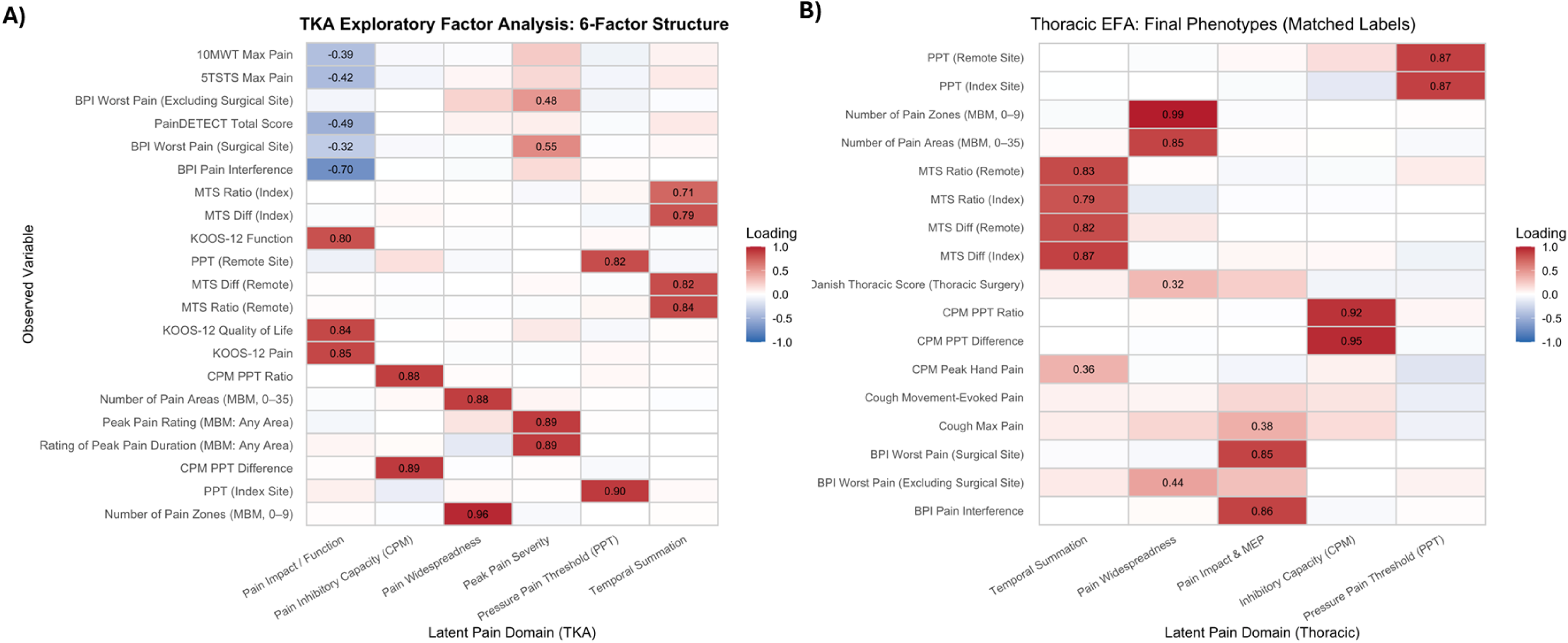
Summary of exploratory factor analysis using pain and pain sensitivity metrics specific to each cohort indicating a 6-factor structure in the A) knee cohort and a 4-factor structure in the B) thoracic cohort. 10MWT = 10 meter walk test, 5TSTS = Five Times Sit-to-Stand, BPI = Brief Pain Inventory, MTS = mechanical temporal summation, MBM = Michigan Body Map, Diff = difference score, KOOS-12 = Knee injury and Osteoarthritis Outcome Score-12, PPT = pressure pain thresholds, CPM = conditioned pain modulation.

### Network Analysis

The network analysis reveals an organized pattern of interrelationships that illustrates how pain and pain sensitivity assessments correlate and jointly characterize multiple dimensions of the pain experience. In the combined cohort, all measures except CPM metrics are well connected in the network, indicating strong connectivity with multiple other measures and suggesting shared variance across multiple pain dimensions (see **Figure 5** and link to dynamic visualization in **Supplemental Materials II**). However, this structure varied across cohorts. In the knee cohort, self-report measures of peak pain severity, pain widespreadness, and pain impact form a strongly correlated cluster, while the QST pain sensitivity measures emerged as relatively isolated, with few correlations exceeding the linkage threshold (**Figure 6a**). In contrast, the thoracic cohort showed weaker associations among self-report measures across most domains, with CPM metrics appearing isolated, indicating inhibitory function is a distinct and largely independent domain within this cohort (**Figure 6b**).

**Figure 5.**
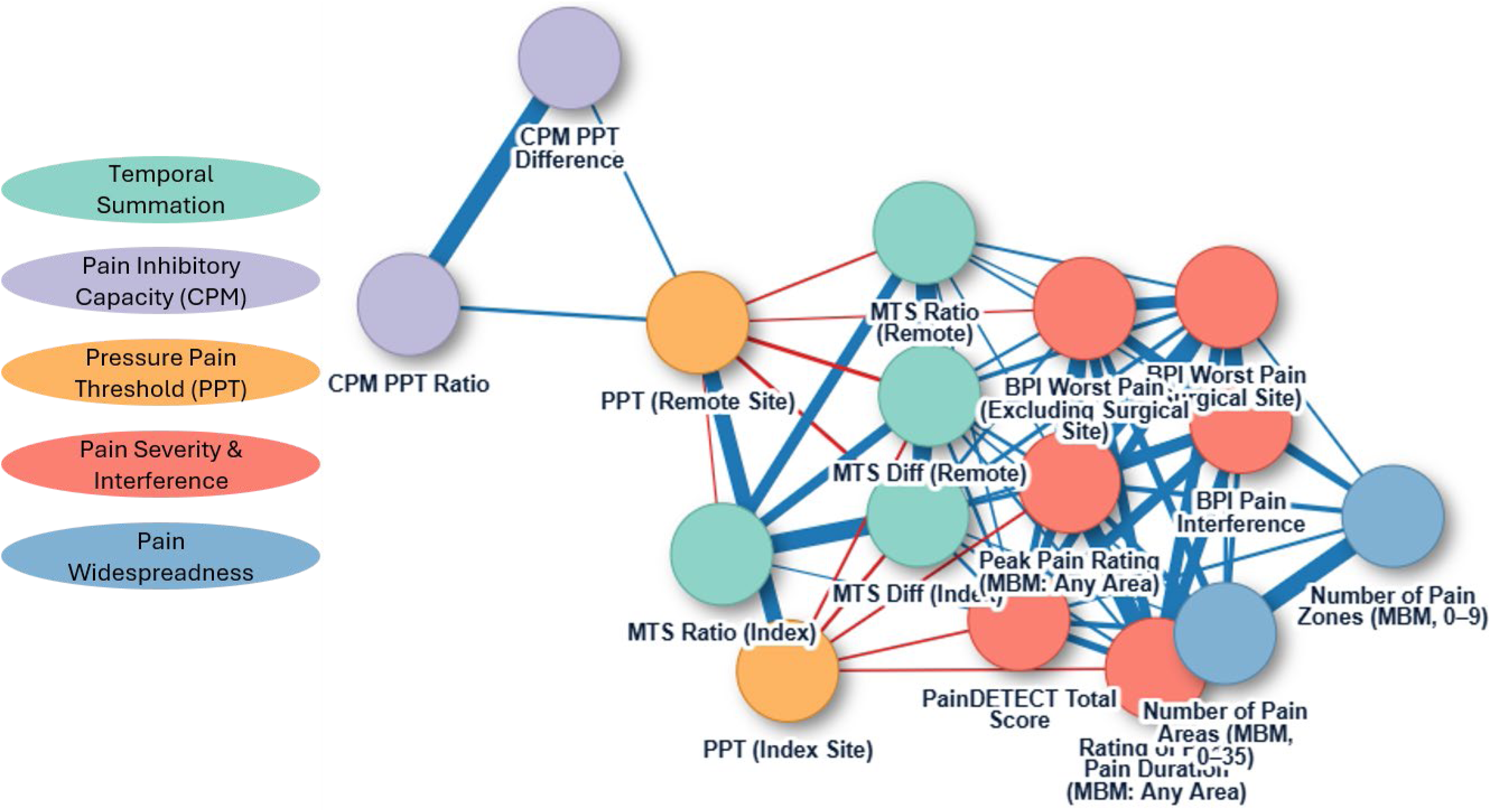
Network analysis indicating edges (lines) defining correlations between nodes (pain metrics) using data from both cohorts. Node color designates latent domains established from exploratory factor analysis. CPM = conditioned pain modulation, PPT = pressure pain threshold, MTS = mechanical temporal summation, BPI = Brief Pain Inventory, Diff = difference score, MBM = Michigan Body Map. Link to interactive network analysis: <u>file:///C:/Users/berard/AppData/Local/Microsoft/Olk/Attachments/ooa-2e815ea8-569f-404c-97d9-0955eb91bf67/304a701909cab5fc541fe46d1734e9aa0172bd1635288d11d07a4e41be2ec2a4/factor_analysis_7_17_26.html</u>

**Figure 6.**
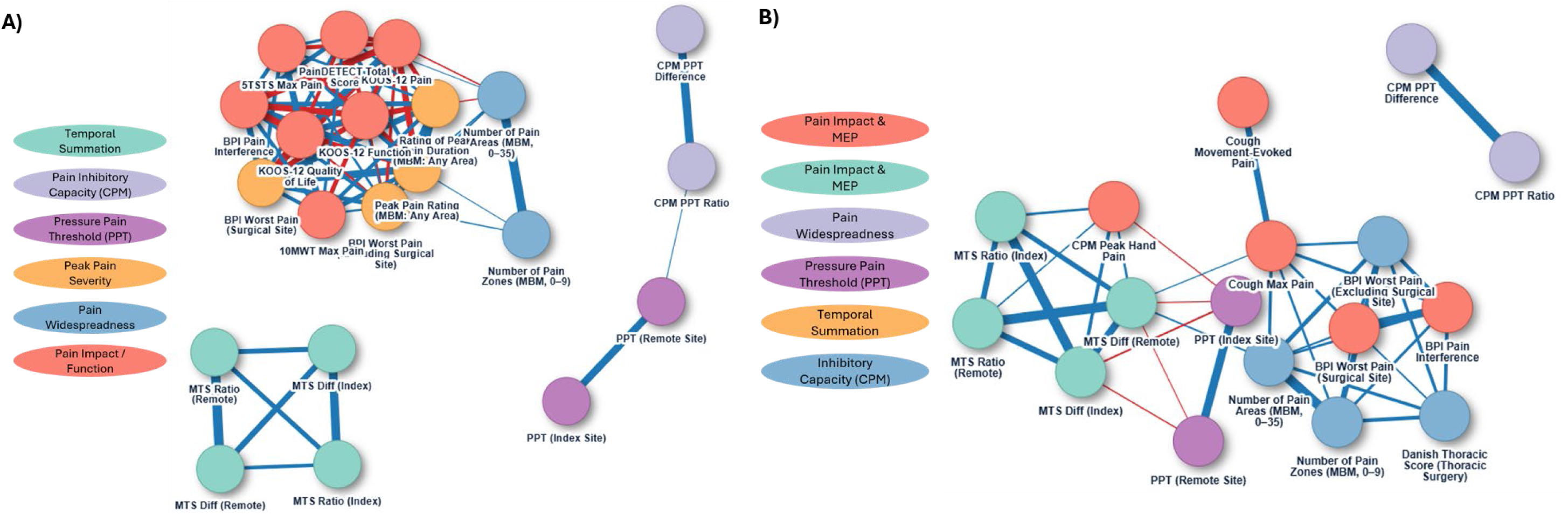
Network analysis indicating edges (lines) defining correlations between nodes (pain metrics) using data from the A) knee cohort and B) thoracic cohort. Node color designates latent domains established from exploratory factor analysis. 5TSTS = Five Times Sit-to-Stand, 10MWT = 10 meter walk test, KOOS-12 = Knee injury and Osteoarthritis Outcome Score-12, CPM = conditioned pain modulation, PPT = pressure pain threshold, MTS = mechanical temporal summation, BPI = Brief Pain Inventory, Diff = difference score, MBM = Michigan Body Map. Link to interactive network analysis: <u>file:///C:/Users/berard/AppData/Local/Microsoft/Olk/Attachments/ooa-2e815ea8-569f-404c-97d9-0955eb91bf67/304a701909cab5fc541fe46d1734e9aa0172bd1635288d11d07a4e41be2ec2a4/factor_analysis_7_17_26.html</u>

## Discussion

The A2CPS program implemented a harmonized, multidimensional battery of pain-related assessments spanning self-report, QST, movement⍰evoked pain, and daily symptom trajectories. This manuscript consolidates the pain assessment battery and clarifies how the baseline measures relate to one another. Using descriptive statistics, correlation heat maps, exploratory factor analyses, and dynamic network/linkage models, we show that the dataset consistently exhibits a multidimensional structure: five robust pain and pain sensitivity domains—pain inhibitory capacity, pain severity and interference, pain widespreadness, pressure pain threshold, and temporal summation, emerged across the full cohort and reproduced in both knee and thoracic cohorts with similar loading patterns. This was observed even though knee participants entered the study with substantial baseline pain while thoracic participants reported minimal thoracic pain, potentially resulting in the separation of pain severity from interference in the knee cohort. Together, these results argue against any single index of “pain” and instead support multifactorial constructs—e.g., intensity/interference indexing subjective burden, TS/CPM indexing facilitation and inhibition, cuff pain and TS capturing temporal dynamics under sustained input, and MEP isolating function⍰relevant pain. These results provide the research community with a clear empirical framework for variable selection and modeling for continued and future use of the full A2CPS dataset and its interpretation.

### Interpretation in context of pain mechanisms

The pattern of higher surgical-site pain and interference in the knee cohort, together with greater temporal summation (both locally and at a remote site), aligns with a profile of augmented central pain facilitation in osteoarthritis-related pain prior to surgery^77,78^. In both cohorts, average CPM values reflected net inhibition, but with small effect sizes, suggesting attenuated descending inhibition, at least in subgroups. This phenotype has been linked to greater acute post-operative pain and increased risk for chronic postsurgical pain in various clinical populations^68–70,79,80^. These findings support a mechanism-informed view where individuals enter surgery with different balances of facilitatory and inhibitory pain processing, differences that may meaningfully influence acute recovery and chronic pain risk. The broadly comparable remote-site PPTs suggest shared baseline pressure sensitivity across cohorts, whereas the divergent TS and cuff pain responses point to condition-specific sensitization profiles — a distinction that may reflect the significant chronic pain burden inherent to degenerative joint disease but minimal in many thoracic surgical candidates at time of enrollment.

Widespread pain and pain lasting longer than 6 months at any location were reported in both cohorts, including thoracic participants who had low index-site pain, but frequently reported other areas of pain. This suggests that many surgical candidates present with multiple concurrent pain conditions, which may or may not be related to the planned surgical procedure. Both widespread pain and long-standing symptoms have been associated with impaired function, slower post⍰operative recovery, and increased likelihood of chronic pain^55,81–83^. Incorporating these features into predictive modeling may improve identification of individuals at elevated risk and help support tailored perioperative management strategies, which are central to the original aims of the A2CPS program.

### Sex differences

Although sex differences were not a primary aim of this study, observed patterns were consistent with the broader literature showing that women tend to report higher pain burden across domains^84–89^. Women in the knee cohort reported higher pain intensity, pain interference, and neuropathic-like symptoms than men, while sex differences in the thoracic cohort were smaller but evident for several pain sensitivity and widespread pain characteristics. These findings underscore the importance of considering sex as a contextual factor when interpreting pain⍰related measures within the A2CPS dataset. Future work may explore whether these differences reflect measurement characteristics or underlying mechanistic features.

### Multifactorial/Comprehensive Pain-Related Variables

A strength of A2CPS is the breadth and depth of its pain-related assessments, collected from pre-operative baseline through the peri-operative period and out to the 6-month (primary endpoint) and 12-month follow-ups. The five domains—pain inhibitory capacity, pain severity and interference, widespreadness, pressure pain threshold, and temporal summation represent distinct physiological and experiential components of the pain phenotype. These findings support the multidimensional nature of pain and reinforce the value of assessing multiple complementary metrics rather than relying on any single measure. While this manuscript focuses on pain and pain sensitivity measures, the broader dataset includes additional psychosocial, behavioral, imaging, and biological “omics” variables. For example, assessments range from pain catastrophizing, fear of movement, anxiety, depression, trauma history, resilience, fatigue, functional ability, activity levels, multisensory sensitivity, opioid use and misuse, genetics, metabolomics, inflammation markers, imaging metrics, etc. These resources create a uniquely comprehensive platform for examining interactions between pain and other multifactorial domains and for conducting innovative secondary analyses as additional timepoints and subjects become publicly available.

### Limitations

Interpretation should consider several design and measurement constraints. First, DMA assessment exhibited floor effects at baseline—most participants reported no pain to light tactile stimulation—yielding near⍰zero variance and limiting inclusion in multivariate models. This is unsurprising given that enrollment preceded thoracic surgery, when allodynia would not yet be expected. Post-operative DMA assessments may yield greater variability and warrant examination in future analyses. Second, cuff⍰evoked pain measures were available only for participants eligible for and completing the fMRI sub-study, so any reason for imaging ineligibility or nonparticipation resulted in systematic missingness for cuff variables; per our pre-specified PRISMA rules, cuff items exceeding the missingness threshold were excluded from EFA. Third, cohort-specific instruments and variable availability (e.g., KOOS⍰12 and movement-evoked pain tasks in the knee cohort; thoracic⍰specific symptom measures in the thoracic cohort), together with the smaller thoracic sample size, constrain cross⍰cohort comparisons and reduce factor stability in that group. Finally, these analyses reflect baseline, cross⍰sectional data; postoperative outcomes were not examined here.

## Conclusions

The A2CPS demonstrates that a rigorous, multidimensional pain assessment can be implemented at scale and sustained across two large surgical cohorts. By documenting the acquisition and quality of these pain and pain sensitivity measures and characterizing the inter-relationships among baseline pain metrics, this manuscript provides a foundation for understanding the empirical structure of the available A2CPS pain battery. The combined self-report pain intensity and interference, widespread pain mapping, QST domains (PPT, TS, CPM, DMA), cuff-evoked responses, daily pain trajectories, and movement-evoked pain offer complementary and only partial overlapping perspectives of pain burden and mechanisms. Baseline differences between cohorts and sexes, together with modest cross⍰domain correlations, underscore the need for mechanism⍰informed, multimodal models rather than reliance on single or isolated pain measures to predict—and ultimately prevent—chronic postsurgical pain. This dataset, supported by rigorous training, data monitoring, and transparent data handling, provides a robust and well curated platform for biomarker discovery and for investigators to leverage in future analyses.

## Data Availability

The data that support the findings of this study are from the Acute to Chronic Pain Signatures (A2CPS) Consortium Data Release 2.1.0.

https://healdata.org/portal/discovery/HDP01069/

## Author Contributions

All authors have reviewed and approved the manuscript.

## Competing Interests

The authors declare that this research was conducted without any commercial, financial, consultant, or institutional relationships that could be construed as a potential conflict of interest.

## Source of Funding

The A2CPS Consortium is supported by the National Institutes of Health Common Fund, which is managed by the OD/Office of Strategic Coordination (OSC). Consortium components include: Clinical Coordinating Center (U24NS112873), Data Integration and Resource Center (U54DA049110), Omics Data Generation Centers (U54DA049116, U54DA049113, U54DA049115), and Multisite Clinical Centers: MCC 1 (UM1NS112874) and MCC 2 (UM1NS118922). Postdoctoral support for GB provided by the National Institutes of Neurological Disease and Stroke (U24NS112873-03S2).

## Notes

### Competing Interest Statement

The authors have declared no competing interest.

### Clinical Protocols

https://pubmed.ncbi.nlm.nih.gov/35547202/

### Author Declarations

All participants provided electronic informed consent as approved by the central Institutional Review Board housed at the University of Iowa.

