## Supplemental Tables and Figures for "Pain and Pain Sensitivity Assessments in the Acute to Chronic Pain Signatures (A2CPS) Program"

**Supplemental Materials**

**Figure S2**. Pearson correlation heat map for A) males and B) females in the knee arthroplasty cohort, showing which pain and pain sensitivity metrics most strongly correlate positively (dark blue) or negatively (dark red), with minimal correlation between metrics represented as white. Note the overall similarity in the heat maps between males and females. BPI = Brief Pain Inventory, nonSS = non-surgical site, MBMc = Michigan Body Map chronic, SS = surgical site, PDQ = PainDETECT Questionnaire, P4 Cuff = individualized pain 4/10 cuff condition, 120 Cuff = standardized 120mmHg cuff condition, TS = temporal summation, Diff = difference score, PPT = pressure pain threshold, MTS = mechanical temporal summation, Shld = shoulder, CPM = conditioned pain modulation, Dur = duration, KOOS-12 = Knee injury and Osteoarthritis Outcome Score-12, QOL = Quality of life, 10MWT = 10 meter walk test, MEP = movement-evoked pain, 5TSTS = Five Times Sit-to-Stand........ 9

**Figure S3**. Pearson correlation heat map for A) males and B) females in the thoracic surgery cohort, showing which pain and pain sensitivity metrics most strongly correlate positively (dark blue) or negatively (dark red), with minimal correlation between metrics represented as white. Note the overall similarity in the heat maps between males and females. BPI = Brief Pain Inventory, nonSS = non-surgical site, MBMc = Michigan Body Map chronic, SS = surgical site, PDQ = PainDETECT Questionnaire, P4 Cuff = individualized pain 4/10 cuff condition, 120 Cuff = standardized 120mmHg cuff condition, TS = temporal summation, Diff = difference score, PPT = pressure pain threshold, MTS = mechanical temporal summation, Shld = shoulder, CPM = conditioned pain modulation, Dur = duration, KOOS-12 = Knee injury and Osteoarthritis Outcome Score-12, QOL = Quality of life, 10MWT = 10 meter walk test, MEP = movement-evoked pain, 5TSTS = Five Times Sit-to-Stand, Thor = thoracic, Cont = control site, DTSQ = Danish Thoracic Surgery Questionnaire, DMA = dynamic mechanical allodynia. ........ 11

**Pain and Pain Sensitivity Assessments in the Acute to Chronic Pain Signatures (A2CPS) Program**

**Supplemental Materials**

**Table S1**. Knee Participant Characteristics & Self-Report Pain Measures by Sex

| **Characteristic** | **Male**<br>N = 381[1] | **Female**<br>N = 649[1] | **p-value**[2] |
|---|---|---|---|
| **Age** | 65.54 (8.29) [377] | 65.03 (8.55) [640] | 0.3 |
| **BPI PI** | 3.79 (2.49) [356] | 4.64 (2.46) [613] | <0.001 |
| **BPI Worst Pain SS** | 5.78 (2.65) [360] | 6.40 (2.52) [614] | <0.001 |
| **BPI Worst Pain exSS** | 3.40 (2.78) [356] | 4.50 (2.93) [610] | <0.001 |
| **MBMc # Pain Areas** | 2.80 (2.91) [363] | 3.75 (4.06) [628] | <0.001 |
| **MBMc # Pain Zones** | 2.04 (1.63) [363] | 2.32 (1.84) [628] | 0.013 |
| **MBMc Peak Pain Rate-Any** | 5.21 (2.37) [267] | 6.08 (2.36) [483] | <0.001 |
| **MBMc Peak Pain Rate Dur** | | | 0.4 |
| **<6mo** | 29 / 254 (11%) | 63 / 470 (13%) | |
| **>=6mo** | 225 / 254 (89%) | 407 / 470 (87%) | |
| **MBMc Peak Pain Dur-Any** | | | >0.9 |
| **<6mo** | 17 / 255 (6.7%) | 31 / 474 (6.5%) | |
| **>=6mo** | 238 / 255 (93%) | 443 / 474 (93%) | |
| **MBMc Peak Pain Dur Rate** | 4.54 (2.34) [254] | 5.16 (2.54) [468] | 0.001 |
| **MBMc Peak Pain Dur-Knee** | | | 0.5 |
| | 18 / 246 (7.3%) | 40 / 464 (8.6%) | |
| | 228 / 246 (93%) | 424 / 464 (91%) | |
| **MBMc Peak Pain Dur Rate-Knee** | 5.00 (2.43) [244] | 5.49 (2.49) [458] | 0.011 |
| **PDQ Total Score** | 7.27 (5.88) [344] | 8.14 (6.54) [598] | 0.036 |
| **KOOS-12 Pain** | 44.42 (16.80) [374] | 39.69 (16.71) [638] | <0.001 |
| **KOOS-12 Function** | 55.56 (21.40) [375] | 49.22 (21.06) [638] | <0.001 |
| **KOOS-12 QOL** | 32.87 (17.68) [374] | 28.98 (17.63) [639] | <0.001 |
| **KOOS-12 Summary** | 44.28 (16.70) [373] | 39.32 (16.19) [637] | <0.001 |

SS = surgical site, nonSS = non-surgical site, BPI = Brief Pain Inventory, MBMc = Michigan Body Map Chronic, dur = duration, PDQ = PainDETECT Questionnaire, KOO-12 = Knee injury and Osteoarthritis Outcome Score-12, DTSQ = Danish Thoracic Surgery Questionnaire, QOL = Quality of life

1 Mean (SD) [n]; n / N (%)

2 Welch Two Sample t-test; Pearson’s Chi-squared test

**Table S2**. Knee Cohort Quantitative Sensory Testing Measures by Sex

| Characteristic | Male<br>N = 381[1] | Female<br>N = 649[1] | p-value[2] |
|---|---|---|---|
| **PPT Index (PPT Knee)** | 3.47 (1.88) [374] | 2.38 (1.37) [633] | <0.001 |
| **PPT Shld** | 4.11 (1.99) [376] | 2.70 (1.36) [633] | <0.001 |
| **MTS Index Diff (MTS Knee Diff)** | 2.22 (1.71) [368] | 2.44 (1.84) [608] | 0.057 |
| **MTS Index Ratio (MTS Knee Ratio)** | 2.19 (1.05) [368] | 2.34 (1.24) [608] | 0.044 |
| **MTS Shld Diff** | 1.61 (1.36) [374] | 1.63 (1.58) [621] | 0.9 |
| **MTS Shld Ratio** | 2.07 (0.98) [374] | 2.14 (1.22) [621] | 0.4 |
| **CPM Ratio** | -5.40 (29.84) [365] | -8.52 (36.81) [611] | 0.15 |
| **CPM Diff** | -0.12 (1.14) [365] | -0.11 (0.79) [611] | 0.9 |
| **CPM Peak Hand Pain** | 5.63 (2.91) [369] | 6.23 (3.00) [626] | 0.002 |
| **P4 Cuff Pain Begin** | 3.44 (1.85) [206] | 3.25 (2.14) [345] | 0.3 |
| **120 Cuff Pain Begin** | 2.73 (1.86) [194] | 3.12 (2.32) [281] | 0.048 |
| **P4 Cuff TS Diff** | 1.52 (1.99) [205] | 1.87 (2.23) [343] | 0.054 |
| **120 Cuff TS Diff** | 0.97 (1.87) [192] | 1.50 (2.04) [280] | 0.004 |
| **P4 Cuff Pressure** | 157.50 (60.26) [272] | 124.99 (47.12) [453] | <0.001 |

PPT = pressure pain threshold, MTS = mechanical temporal summation, CPM = conditioned pain modulation, P4 Cuff = individualized pain 4/10 cuff condition, 120 Cuff = standardized 120mmHg cuff condition

1 Mean (SD); n / N (%)

2 Welch Two Sample t-test; Pearson’s Chi-squared test

**Table S3**. Knee Cohort Pain with Activity Measures by Sex

| **Characteristic** | **Male** N = 381[1] | **Female** N = 649[1] | **p-value**[2] |
|---|---|---|---|
| **5TSTS Pain Max** | 3.44 (2.49) [369] | 3.81 (2.68) [612] | 0.027 |
| **5TSTS MEP** | 1.81 (1.84) [369] | 1.87 (2.06) [611] | 0.6 |
| **10MWT Pain Max** | 2.83 (2.39) [374] | 3.28 (2.63) [632] | 0.006 |
| **10MWT MEP** | 0.71 (1.17) [372] | 0.63 (1.34) [631] | 0.3 |

5TSTS = Five times sit to stand test; 10MWT = 10-meter walk test; Pain Max = Peak pain with movement; MEP = movement-evoked pain (difference between max and initial resting pain).

1 Mean (SD); n / N (%)

2 Welch Two Sample t-test; Pearson’s Chi-squared test

**Table S4**. Thoracic Participant Characteristics & Self-Report Pain Measures by Sex

| Characteristic | Male N = 158[1] | Female N = 198[1] | p-value[2] |
|---|---|---|---|
| **Age** | 60.97 (12.45) [153] | 58.92 (13.35) [194] | 0.14 |
| **BPI PI** | 0.95 (1.99) [144] | 1.06 (1.98) [176] | 0.6 |
| **BPI Worst Pain SS** | 0.80 (1.89) [151] | 1.15 (2.39) [183] | 0.13 |
| **BPI Worst Pain exSS** | 1.61 (2.47) [144] | 2.67 (3.01) [175] | <0.001 |
| **MBMc # Pain Areas** | 2.71 (3.34) [153] | 3.68 (4.51) [185] | 0.024 |
| **MBMc # Pain Zones** | 1.59 (1.97) [153] | 2.14 (2.30) [185] | 0.021 |
| **MBMc Peak Pain Rate-Any** | 4.07 (2.40) [68] | 5.29 (2.55) [95] | 0.002 |
| **MBMc Peak Pain Rate Dur** | | | 0.007 |
| **<6mo** | 18 / 59 (31%) | 11 / 88 (13%) | |
| **>=6mo** | 41 / 59 (69%) | 77 / 88 (88%) | |
| **MBMc Peak Pain Dur-Any** | | | 0.002 |
| **<6mo** | 11 / 59 (19%) | 3 / 88 (3.4%) | |
| **>=6mo** | 48 / 59 (81%) | 85 / 88 (97%) | |
| **MBMc Peak Pain Dur Rate** | 3.37 (2.25) [59] | 4.32 (2.40) [88] | 0.017 |
| **PDQ Total Score** | 3.73 (5.14) [48] | 6.21 (6.58) [61] | 0.029 |
| **DTS Score** | 11.22 (12.53) [148] | 16.24 (13.75) [186] | <0.001 |

SS = surgical site, nonSS = non-surgical site, BPI = Brief Pain Inventory, MBMc = Michigan Body Map Chronic, dur = duration, PDQ = PainDETECT Questionnaire, KOO-12 = Knee injury and Osteoarthritis Outcome Score-12, DTSQ = Danish Thoracic Surgery Questionnaire, QOL = Quality of life

1 Mean (SD) [n]; n / N (%)

2 Welch Two Sample t-test; Pearson’s Chi-squared test

**Table S5**. Thoracic Cohort Quantitative Sensory Testing Measures by Sex

| Characteristic | Male<br>N = 158[1] | Female<br>N = 198[1] | p-value[2] |
|---|---|---|---|
| **PPT Index (PPT Thor / PPT Knee)** | 2.70 (1.30) [156] | 1.99 (1.36) [190] | <0.001 |
| **PPT Shld** | 3.99 (1.84) [156] | 2.78 (1.64) [190] | <0.001 |
| **MTS Diff (MTS Thor Diff / MTS Knee Diff)** | 1.56 (1.44) [156] | 1.77 (1.80) [187] | 0.2 |
| **MTS Ratio (MTS Thor Ratio / MTS Knee Ratio)** | 2.09 (1.00) [156] | 2.22 (1.44) [187] | 0.3 |
| **MTS Shld Diff** | 0.99 (0.87) [155] | 1.22 (1.36) [188] | 0.059 |
| **MTS Shld Ratio** | 1.80 (0.70) [155] | 1.87 (1.00) [188] | 0.4 |
| **CPM Ratio** | -6.16 (25.89) [150] | -16.47 (41.50) [182] | 0.006 |
| **CPM Diff** | -0.16 (0.92) [150] | -0.38 (0.96) [182] | 0.039 |
| **CPM Peak Hand Pain** | 6.17 (2.79) [150] | 6.69 (2.83) [182] | 0.092 |
| **P4 Cuff Pain Begin** | 3.35 (1.72) [73] | 3.07 (2.06) [96] | 0.3 |
| **120 Cuff Pain Begin** | 2.63 (2.08) [71] | 2.59 (2.18) [85] | 0.9 |
| **P4 Cuff TS Diff** | 1.34 (1.61) [73] | 1.91 (2.44) [95] | 0.074 |
| **120 Cuff TS Diff** | 0.51 (1.63) [71] | 1.21 (2.19) [85] | 0.023 |
| **P4 Cuff Pressure** | 175.91 (65.10) [115] | 155.52 (61.69) [145] | 0.011 |
| **DMA Control Site** | 0.00 (0.01) [155] | 0.01 (0.07) [190] | 0.3 |
| **DMA Index Site** | 0.00 (0.01) [156] | 0.01 (0.10) [190] | 0.2 |

PPT = pressure pain threshold, Shld = shoulder, MTS = mechanical temporal summation, CPM = conditioned pain modulation, P4 Cuff = individualized pain 4/10 cuff condition, 120 Cuff = standardized 120mmHg cuff condition, DMA = dynamic mechanical allodynia

1 Mean (SD); n / N (%)

2 Welch Two Sample t-test; Pearson’s Chi-squared test

**Table S6**. Thoracic Cohort Pain with Activity Measures by Sex

| Characteristic | Male<br>N = 158[1] | Female<br>N = 198[1] | p-value[2] |
|---|---|---|---|
| Deep Breathing Max Pain | 0.33 (1.04) [158] | 0.42 (1.14) [196] | 0.4 |
| Coughing Max Pain | 0.46 (1.37) [158] | 0.63 (1.36) [196] | 0.3 |
| Deep Breathing MEP | 0.03 (0.43) [158] | 0.07 (0.69) [196] | 0.6 |
| Coughing MEP | 0.16 (0.93) [158] | 0.27 (0.83) [196] | 0.3 |

MEP = movement-evoked pain

1 Mean (SD); n / N (%)

2 Welch Two Sample t-test; Pearson’s Chi-squared test

**Pain and Pain Sensitivity Assessments in the Acute to Chronic Pain Signatures (A2CPS) Program**

**Supplemental Materials**

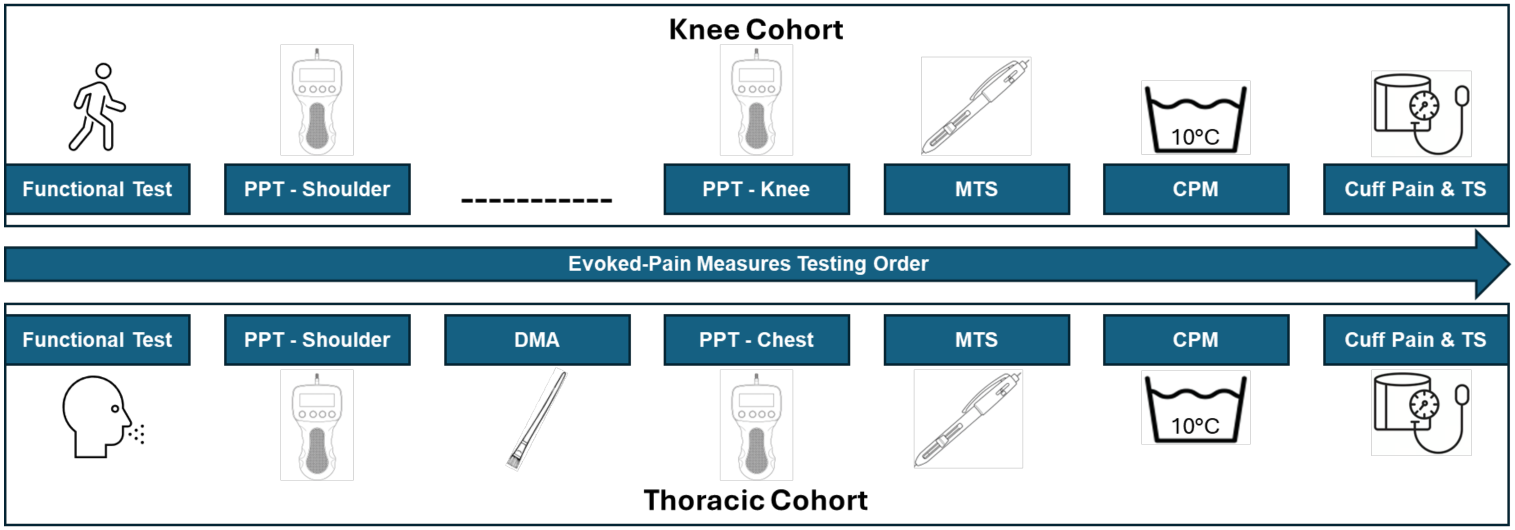

**Figure S1**. Order of evoked-pain measures for the knee (top) and thoracic (bottom) cohorts. Note DMA was performed in the thoracic cohort only. PPT = pressure pain sensitivity, DMA = dynamic mechanical allodynia, MTS = mechanical temporal summation, CPM = conditioned pain modulation, TS = temporal summation.

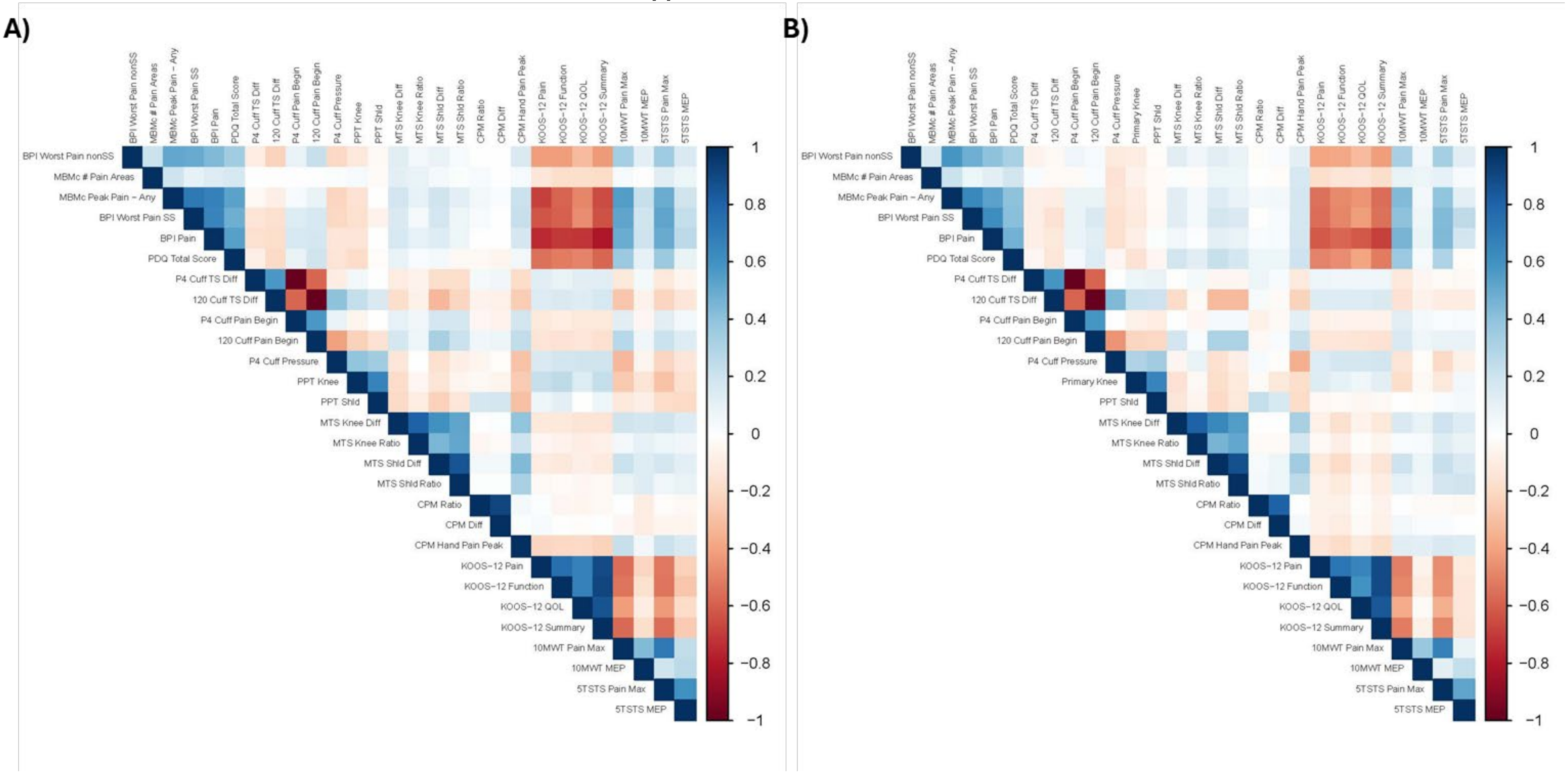

**Figure S2**. Pearson correlation heat map for A) males and B) females in the knee arthroplasty cohort, showing which pain and pain sensitivity metrics most strongly correlate positively (dark blue) or negatively (dark red), with minimal correlation between metrics represented as white. Note the overall similarity in the heat maps between males and females. BPI = Brief Pain Inventory, nonSS = non-surgical site, MBMc = Michigan Body Map chronic, SS = surgical site, PDQ = PainDETECT Questionnaire, P4 Cuff = individualized pain 4/10 cuff condition, 120 Cuff = standardized 120mmHg cuff condition, TS = temporal summation, Diff = difference score, PPT = pressure pain threshold, MTS = mechanical temporal summation, Shld = shoulder, CPM = conditioned pain modulation, Dur = duration, KOOS-12 = Knee injury and Osteoarthritis Outcome Score-12, QOL = Quality of life, 10MWT = 10 meter walk test, MEP = movement-evoked pain, 5TSTS = Five Times Sit-to-Stand.

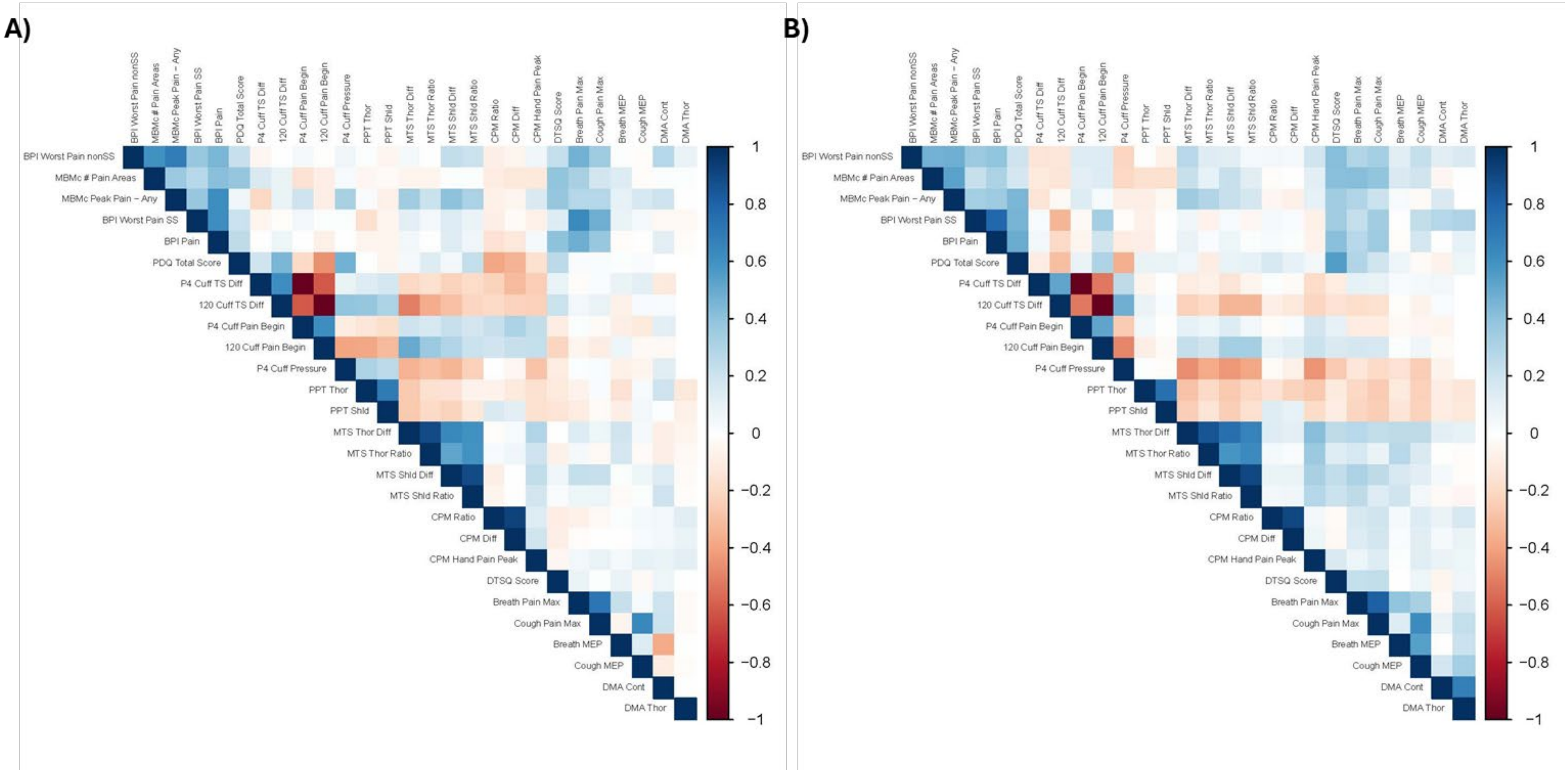

**Figure S3**. Pearson correlation heat map for A) males and B) females in the thoracic surgery cohort, showing which pain and pain sensitivity metrics most strongly correlate positively (dark blue) or negatively (dark red), with minimal correlation between metrics represented as white. Note the overall similarity in the heat maps between males and females. BPI = Brief Pain Inventory, nonSS = non-surgical site, MBMc = Michigan Body Map chronic, SS = surgical site, PDQ = PainDETECT Questionnaire, P4 Cuff = individualized pain 4/10 cuff condition, 120 Cuff = standardized 120mmHg cuff condition, TS = temporal summation, Diff = difference score, PPT = pressure pain threshold, MTS = mechanical temporal summation, Shld = shoulder, CPM = conditioned pain modulation, Dur = duration, KOOS-12 = Knee injury and Osteoarthritis Outcome Score-12, QOL = Quality of life, 10MWT = 10 meter walk test, MEP = movement-evoked pain, 5TSTS = Five Times Sit-to-Stand, Thor = thoracic, Cont = control site, DTSQ = Danish Thoracic Surgery Questionnaire, DMA = dynamic mechanical allodynia.

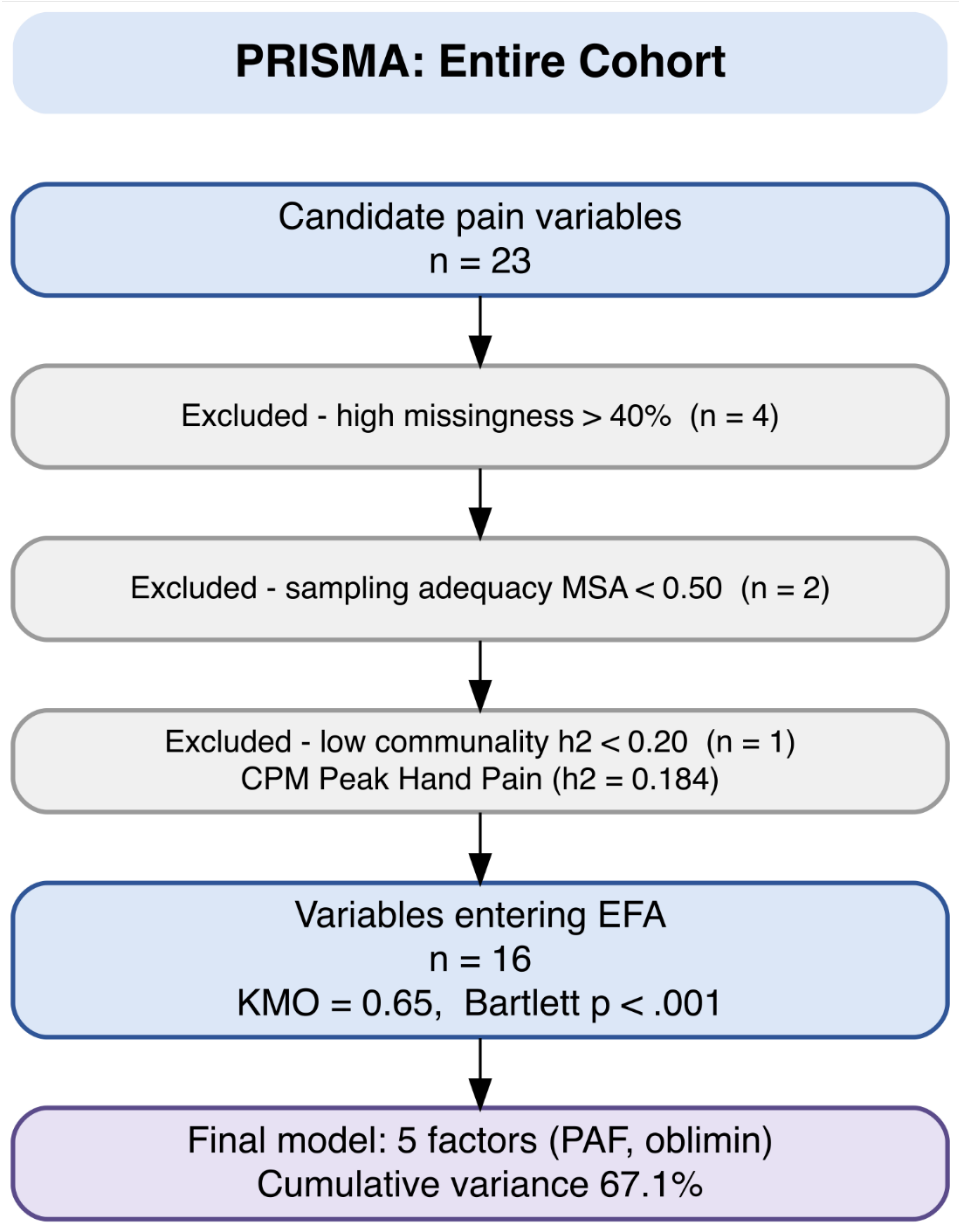

**Figure S4**. PRISMA flow diagram outlining the process used to identify variable selection in the exploratory factor analysis for the entire cohort (knee plus thoracic patients). MSA = measure of sample adequacy, CPM = conditioned pain modulation, EFA = exploratory factor analysis, KMO = Kaiser-Meyer-Olkin test, PAF = principal axis factoring.

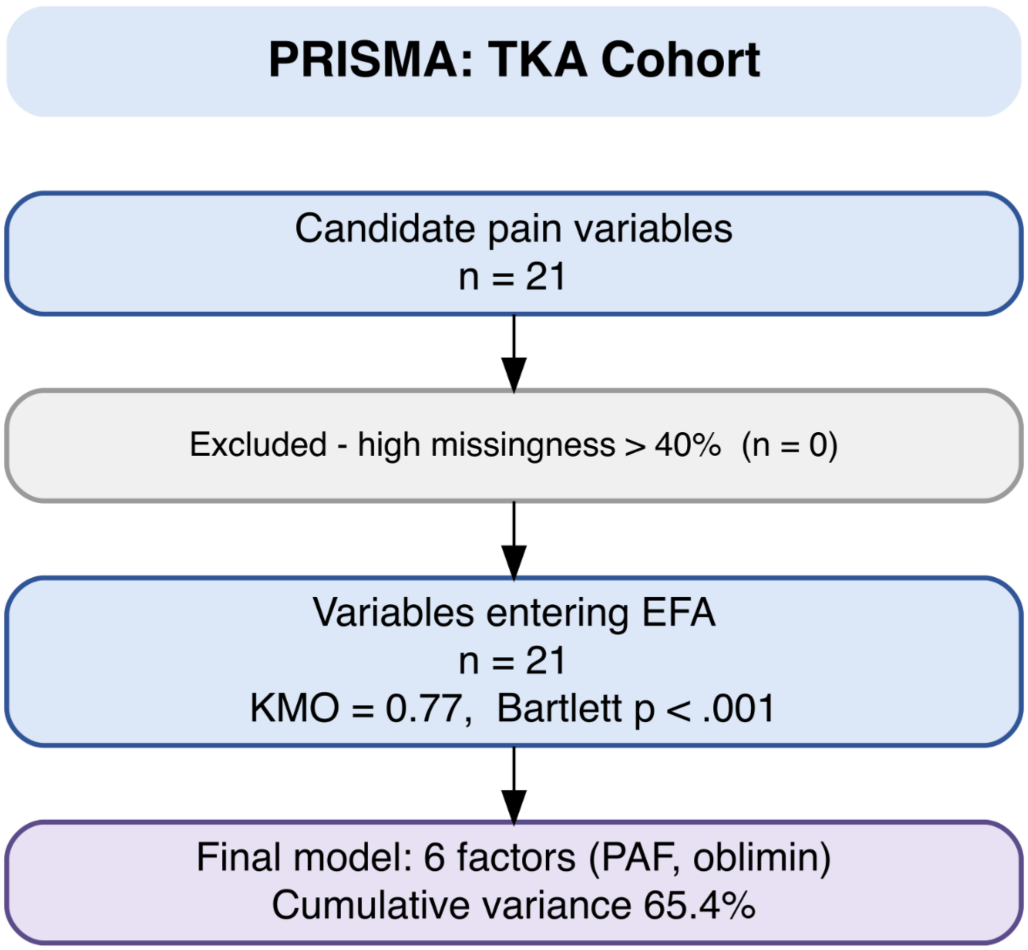

**Figure S5**. PRISMA flow diagram outlining the process used to identify variable selection in the exploratory factor analysis for the knee (TKA = total knee replacement) cohort. EFA = exploratory factor analysis, KMO = Kaiser-Meyer-Olkin test, PAF = principal axis factoring.

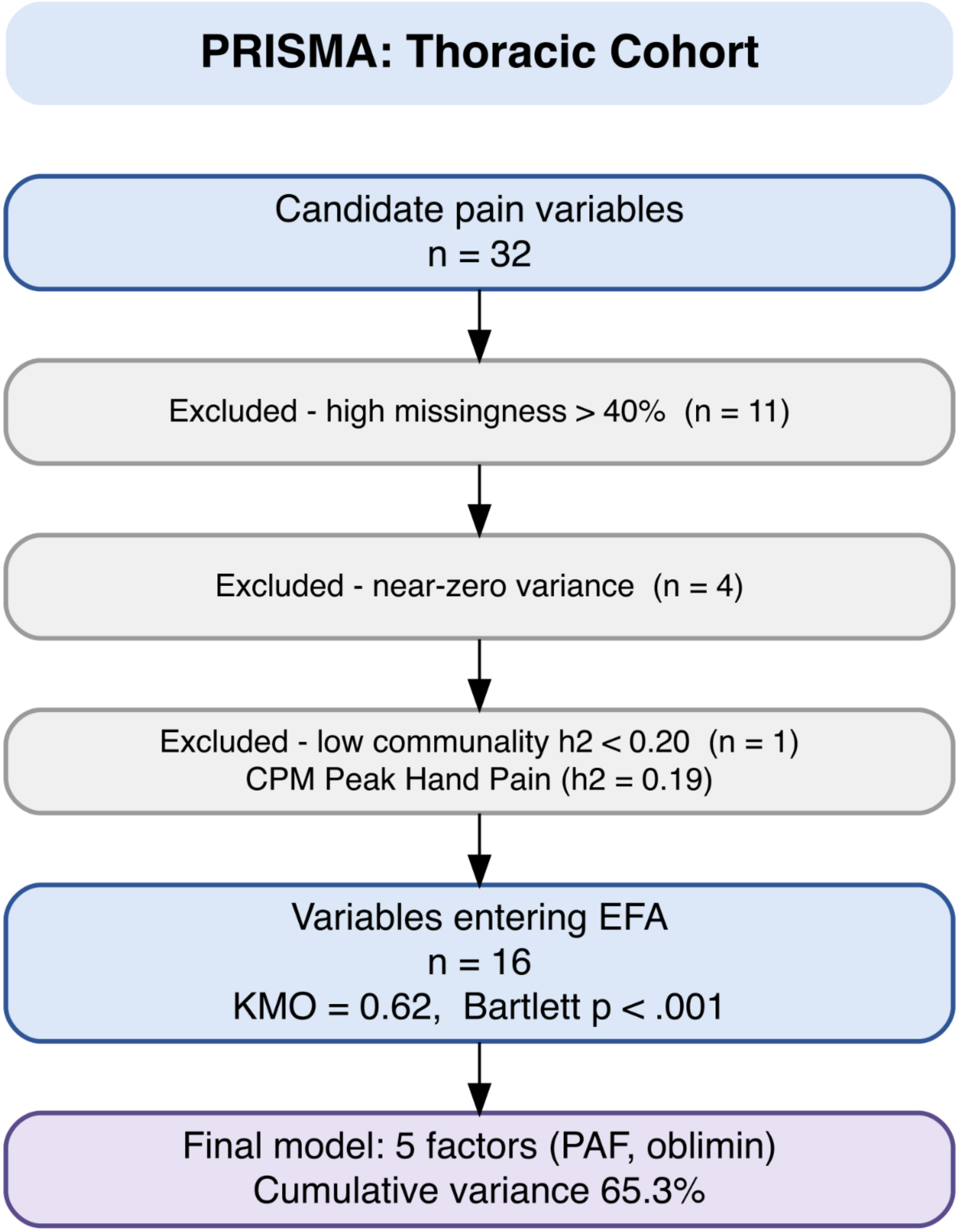

**Figure S6**. PRISMA flow diagram outlining the process used to identify variable selection in the exploratory factor analysis for the thoracic cohort. CPM = conditioned pain modulation, EFA = exploratory factor analysis, KMO = Kaiser-Meyer-Olkin test, PAF = principal axis factoring.
